# Patient-specific forecasting of post-radiotherapy prostate-specific antigen kinetics enables early prediction of biochemical relapse

**DOI:** 10.1101/2022.03.07.22271524

**Authors:** Guillermo Lorenzo, Nadia di Muzio, Chiara Lucrezia Deantoni, Cesare Cozzarini, Andrei Fodor, Alberto Briganti, Francesco Montorsi, Víctor M. Pérez-García, Hector Gomez, Alessandro Reali

## Abstract

The detection of prostate cancer recurrence after external beam radiotherapy relies on the measurement of a sustained rise of serum prostate-specific antigen (PSA). However, this biochemical relapse may take years to occur, thereby delaying the delivery of a secondary treatment to patients with recurring tumors. To address this issue, here we propose to use patient-specific forecasts of PSA dynamics to early predict biochemical relapse. Our forecasts are based on mechanistic models of prostate cancer response to external beam radio-therapy, which are fit to patient-specific PSA data collected during standard post-treatment monitoring. Our results show a remarkable performance of our models in recapitulating the observed changes in PSA and yielding short-term predictions over approximately one year (cohort median RMSE of 0.10 to 0.47 ng/mL and 0.13 to 1.41 ng/mL, respectively). Additionally, we identify three model-based biomarkers that enable an accurate identification of biochemical relapse (AUC *>* 0.80) significantly earlier than standard practice (*p <* 0.01).

## Introduction

External beam radiotherapy (EBRT) is a standard treatment for prostate cancer (PCa) that is potentially available for patients of all ages to treat tumors ranging from low to high and very high risk [Mottet et al., 2021; Wein et al., 2012; Tang et al., 2020; Gray et al., 2017]. During EBRT, the prostate is exposed to an external source of radiation, which aims at disrupting the DNA in tumor cells’ nuclei. The accumulation of radiation-induced damage along with the multiple genetic alterations underlying the development of PCa ultimately forces tumor cells to undergo programmed cell death [Alberts et al., 2007]. EBRT is usually delivered as daily fractions of approximately 2 to 3 Gy until completing a total dose ranging from 60 to 80 Gy. In particular, the use of daily doses higher than 2 Gy in the recent decades has led to so-called moderate hypofractionation [Mottet et al., 2021; Wein et al., 2012]. This treatment modality requires a lower number of radiation sessions, which presents pharmacoeconomic advantages. The efficacy of EBRT can be improved through combination with neoadjuvant or adjuvant androgen deprivation therapy (ADT) [Mottet et al., 2021]. However, since ADT can produce several side effects, it is usually prescribed for intermediate and high-risk PCa patients only (i.e., those who present a higher risk of metastasis).

After the completion of EBRT, patients are monitored using the serum levels of prostate-specific antigen (PSA), which is a standard clinical biomarker of PCa [Mottet et al., 2021; Cornford et al., 2021; Wein et al., 2012]. The rationale for using PSA in post-EBRT patient follow-up is that blood levels of PSA tend to rise due to PCa growth. Thus, if the treatment is successful, radiation-induced tumor cell death should decrease PSA values to a minimum, which may vary from patient to patient and with prostate size [Ray et al., 2006; Roehrborn et al., 2000]. Otherwise, if EBRT does not eradicate the tumor completely, the surviving cancerous cells will ultimately drive tumor growth after EBRT conclusion and, hence, produce an increasing trend in PSA. This phenomenon is termed biochemical relapse and it is thus indicative of tumor recurrence (see Figure 1). Approximately 20% to 50% of PCa patients undergoing radiotherapy as primary curative treatment will ultimately develop biochemical relapse within 5 to 10 years after treatment conclusion, respectively [Kupelian et al., 2006; Rosenbaum et al., 2004]. Following the detection of biochemical relapse, tumor recurrence can be confirmed through biopsy and imaging methods, such as magnetic resonance imaging (MRI) and prostate-specific membrane antigen or choline positron emission tomography/computed tomography (PSMA or choline PET/CT, respectively). To treat post-EBRT PCa recurrence, there are several therapeutic strategies that depend on whether the recurrence is local or metastatic [Cornford et al., 2021; Wein et al., 2012].

**Figure 1.**
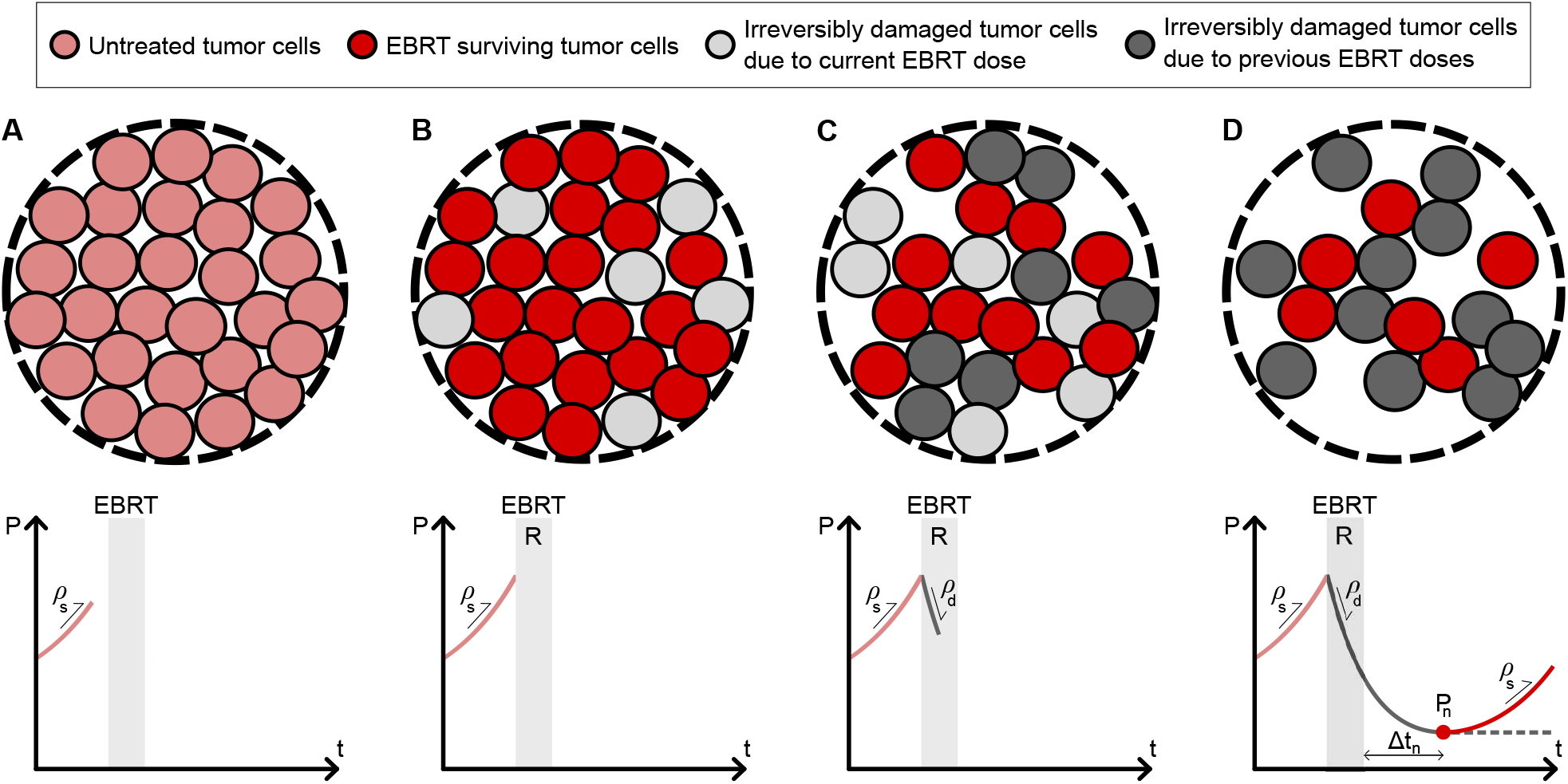
Mechanistic modeling of PSA dynamics after EBRT. This figure illustrates the mechanisms included in our models of PCa response to EBRT, which assume that serum PSA is proportional to the number of cells in the patient’s tumor. The upper row shows the PCa cells in a generic tumor region at four instants before, during, and after EBRT. The bottom row shows the corresponding PSA evolution up to each depicted time instant. (A) Before treatment, we assume that tumor cells grow exponentially at a rate *ρ*_*s*_, which also describes the characteristic increase of PSA in untreated PCa (rose solid line). (B) After the first EBRT dose, a fraction of tumor cells *R* survives to radiation, while the complementary fraction, 1 − *R*, is irreversibly damaged and will ultimately die. This process will repeat with each consecutive EBRT dose. (C) During EBRT, while the surviving cells may continue to proliferate at a rate *ρ*_*s*_, the radiation-damaged cells undergo programmed cell death at a rate *ρ*_*d*_. This becomes the dominant mechanism during EBRT, and produces a decreasing trend in PSA (gray solid line). (D) After the conclusion of EBRT, the remainder of the radiation-damaged cells die and PSA continues to progressively drop. If the treatment does not fully eliminate the tumor, the surviving cells continue proliferating at a rate *ρ*_*s*_ and ultimately produce a biochemical relapse (red solid line). If the treatment eradicates all tumor cells, then PSA reaches a plateau (gray dashed line). Our mechanistic models also enable a quantitative estimation of the PSA nadir (*P*_*n*_) and the time to PSA nadir since EBRT termination (Δ*t*_*n*_). See the STAR Methods for further details on mathematical modeling.

Serum PSA may exhibit natural fluctuations (e.g., due to diet and lifestyle), a smooth increase caused by benign prostatic enlargement (i.e., benign prostatic hyperplasia), and transient peaks due to ADT termination or the so-called PSA bounce, which consists of a temporary PSA increase of 0.1 to 0.5 ng/mL usually occurring within the first 24 months after EBRT conclusion [Wein et al., 2012; Pinkawa et al., 2010; Freiberger et al., 2017; Carobene et al., 2018; Christensson et al., 2011; Roehrborn et al., 2000]. These phenomena may hamper the detection of biochemical relapse following EBRT. Thus, the clinical criteria to identify a biochemical relapse require PSA to exhibit a consistent rising trend over time [Wein et al., 2012; Cornford et al., 2021]. For example, a standard criterion with widespread use in current clinical practice identifies a biochemical relapse as a PSA increase larger than 2 ng/mL over the detected PSA nadir (i.e., the minimum PSA value measured for a patient) [Roach et al., 2006; Cornford et al., 2021]. Additionally, multiple studies have been devoted to analyze PSA dynamics after EBRT and its correlation with the pathological features of tumor recurrence to define further PSA-based markers that improve the identification and prognostic assessment of biochemical relapse and PCa recurrence. For instance, the detection of a rapid decline of PSA right after treatment, overall high PSA values during post-treatment monitoring, an early nadir, a high value of the nadir, and a low PSA doubling time during biochemical relapse (i.e., the time it would take PSA to exhibit a two-fold increase) have been correlated with a poorer prognosis, including metastatic disease and lower patient survival [Freiberger et al., 2017; Zelefsky et al., 2005; Zumsteg et al., 2015; Ray et al., 2006; Bates et al., 2005; Cheung et al., 2006; Cavanaugh et al., 2004; Shi et al., 2013; Wein et al., 2012]. Alternatively, post-EBRT PSA dynamics has also been analyzed by fitting empirical equations to patient-specific longitudinal series of PSA values. In particular, very successful results have been reported by leveraging a biexponential formula, which consists of the sum of two terms: an exponential decay to capture the usual post-treatment decline in PSA observed in all patients and a rising exponential to represent biochemical relapse, which vanishes when this empirical model is fit to data from cured patients [Zagars and Pollack, 1997; Cox et al., 1994; Hanlon et al., 1998; Vollmer and Montana, 1999; Taylor et al., 2005].

However, the current criteria of biochemical relapse and the majority of PSA-based markers only enable to assess this event upon its direct observation. Hence, these approaches may ultimately delay the diagnosis and treatment of tumor recurrence, thereby potentially reducing the chances of successfully controlling the disease. Additionally, observational metrics and models of PSA dynamics offer a limited representation of the underlying tumor dynamics that ultimately regulate the observed changes of PSA in each patient. To address these limitations, we propose to leverage mechanistic models of PCa response to EBRT (see Figure 1) in order to forecast PSA dynamics on a patient-specific basis [Lorenzo et al., 2019b]. Our goal is to use this approach to predict the occurrence of biochemical relapse and, hence, ultimately facilitate an early diagnosis and treatment of tumor recurrence after EBRT. The mechanistic modeling of tumor growth and therapeutic response is an established approach that aims at mathematically describing the biophysical mechanisms underlying these phenomena in order to increase our understanding of cancer diseases and advance their clinical management on a personalized basis [Yankeelov et al., 2013; Rockne et al., 2019; Karolak et al., 2018; Jarrett et al., 2020; Mang et al., 2020; Wang et al., 2009; Lorenzo et al., 2019a; Oden et al., 2016]. In particular, these models can be fit to patient-specific data and then leveraged to render personalized computer forecasts of tumor prognosis and treatment outcomes capable to assist clinical decision-making [Kazerouni et al., 2020; Lorenzo et al., 2021; Mang et al., 2020].

Several studies have investigated mechanistic models of PCa growth and PSA dynamics in various scenarios, including untreated tumor growth [Lorenzo et al., 2016, 2019b; Swanson et al., 2001; Vollmer, 2010; Farhat et al., 2017], hormone therapy [Brady-Nicholls et al., 2021, 2020; Ideta et al., 2008; Hirata et al., 2010; Jain et al., 2011; Morken et al., 2014; Phan et al., 2019; Jackson, 2004], cytotoxic and antiangiogenic therapies [West et al., 2018, 2019; Colli et al., 2020, 2021], and after radical prostatectomy [Vollmer and Humphrey, 2003; Truskinovsky et al., 2005]. Since radiotherapy is used for the treatment of many types of cancer, the study of tumor response to radiation and the forecasting of patient-specific radiotherapeutic outcomes using mechanistic models constitute a rich area of research [Corwin et al., 2013; Hormuth et al., 2021; Rockne et al., 2015; Lipková et al., 2019; Lima et al., 2017; Ayala-Hernández et al., 2021; Pérez-García et al., 2015; Zahid et al., 2021; Alfonso et al., 2021; Powathil et al., 2007]. Nevertheless, there is a dearth of mechanistic models providing a coupled description of tumor and PSA dynamics following radiotherapy [Lorenzo et al., 2019b; Sosa-Marrero et al., 2021; Yamamoto et al., 2016].

In [Lorenzo et al., 2019b], we presented a mechanistic modeling framework to describe how the response of PCa to EBRT drives PSA dynamics after treatment and identified promising model-based biomarkers to detect biochemical relapse. Our modeling framework relies on five key assumptions, which are illustrated in Figure 1. We assume that PCa cells proliferate following an exponential law and that PSA is proportional to the number of tumor cells. We further assume that each radiation dose irreversibly damages a fraction of the tumor cells that ultimately undergoes programmed cell death, whereas the complementary fraction survives to EBRT and continues proliferating. Additionally, we consider that EBRT is delivered either periodically or as an equivalent single dose. This last assumption leads to two alternative models that we have termed as the periodic dose model and the single dose model, respectively (see STAR Methods for further details on mathematical modeling). In the present work, we first validate our models and model-based biomarkers of biochemical relapse from [Lorenzo et al., 2019b] in a new cohort, whose main characteristics are summarized in Table 1 (see STAR Methods for further details). To this end, we perform a global fitting study in which we parameterize our models with all PSA data available for each patient. Then, we assess the predictive performance of our models in a series of fitting-forecasting scenarios. Each scenario leverages an increasing number of the PSA values collected for each patient for model fitting, which is followed by a corresponding personalized model forecast of PSA dynamics that we compare against the remainder of the patient’s PSA data. This approach would simulate the utilization of our models during actual patient monitoring: each newly collected PSA value enables to update the models for a given patient and, hence, obtain an updated prediction of PSA dynamics on an individual basis. Additionally, we analyze the ability of our model-based biomarkers as determined in these fitting-forecasting scenarios to early identify biochemical relapse, and we further assess whether they outperform the standard clinical criteria that were used in this cohort.

**Table 1.** Summary of patient cohort characteristics. All patients received a total of *n*_*d*_ = 28 radiation doses in their EBRT plan. The number of PSA values (*n*_*P*_) includes the baseline PSA (*P*_*d*_). The PSA test frequency refers to post-EBRT monitoring exclusively. IQR: interquartile range.

| Characteristic | All patients ($n=166$) | | | Non-relapsing patients ($n=156$) | | | Relapsing patients ($n=10$) | | |
| --- | --- | --- | --- | --- | --- | --- | --- | --- | --- |
|  | Median | IQR | Range | Median | IQR | Range | Median | IQR | Range |
| **Clinical** |  |  |  |  |  |  |  |  |  |
| Age at diagnosis (y) | 73 | (68, 76) | (49, 83) | 73 | (68, 76) | (49, 83) | 73 | (68, 76) | (65, 79) |
| Baseline PSA, $P_d$ (ng/mL) | 6.3 | (4.6, 8.6) | (0.6, 81.0) | 6.2 | (4.5, 8.4) | (0.6, 22.0) | 9.6 | (5.9, 13.1) | (4.9, 81.0) |
| Gleason score | 6 | (6, 7) | (4, 8) | 6 | (6, 7) | (5, 8) | 7 | (6, 7) | (4, 7) |
| **EBRT** |  |  |  |  |  |  |  |  |  |
| Age at EBRT (y) | 74 | (68, 77) | (49, 84) | 74 | (68, 77) | (49, 84) | 75 | (73, 76) | (66, 80) |
| Total dose (Gy) | 74.2 | (71.4, 74.2) | (65.8, 77.4) | 74.2 | (71.4, 74.2) | (71.4, 77.4) | 74.2 | (71.4, 74.2) | (65.8, 74.2) |
| Dose per fraction (Gy) | 2.65 | (2.55, 2.65) | (2.35, 2.76) | 2.65 | (2.55, 2.65) | (2.55, 2.76) | 2.65 | (2.55, 2.65) | (2.35, 2.65) |
| Duration (mo) | 1.3 | (1.3, 1.4) | (1.2, 2.1) | 1.3 | (1.3, 1.4) | (1.2, 2.1) | 1.3 | (1.3, 1.4) | (1.2, 1.7) |
| **Patient monitoring** |  |  |  |  |  |  |  |  |  |
| No. PSA values, $n_p$ | 11 | (8, 13) | (6, 25) | 11 | (8, 13) | (6, 25) | 14 | (10, 19) | (7, 22) |
| Follow-up since EBRT (y) | 5.7 | (4.5, 7.6) | (3.0, 14.0) | 5.7 | (4.5, 7.5) | (3.0, 14.0) | 5.7 | (4.2, 8.1) | (3.3, 11.2) |
| PSA test frequency (mo) | 6.3 | (4.0, 10.3) | (0.0, 59.4) | 6.4 | (4.1, 11.4) | (0.0, 59.4) | 4.9 | (3.2, 7.3) | (1.0, 20.5) |

## Results

### Mechanistic models recapitulate patient-specific PSA dynamics after EBRT

We begin by performing a global fitting analysis to assess the ability of our mechanistic models in reproducing the longitudinal PSA series collected for each patient in the cohort. To this end, we fit the periodic dose model and the single dose model to all the PSA values available for each patient (see STAR Methods for methodological details). Figure 2 illustrates representative global fitting results for three non-relapsing and three relapsing patients. Using the periodic dose model, the median and interquartile range of the root mean squared error (RMSE) and the coefficient of determination (*R*^2^) of our model fits are 0.17 (0.08, 0.28) ng/mL and 0.99 (0.97, *>* 0.99) over the whole cohort (*n* = 166), 0.15 (0.07, 0.24) ng/mL and 0.99 (0.98, *>* 0.99) in the non-relapsing subgroup (*n* = 156), and 0.53 (0.40, 0.59) ng/mL and 0.95 (0.91, 0.98) in the relapsing subgroup (*n* = 10), respectively. For the single-dose model, the RMSE and *R*^2^ of the model fits are distributed with median and interquartile range of 0.16 (0.07, 0.26) ng/mL and 0.99 (0.98, *>* 0.99) over the whole cohort (*n* = 166), 0.15 (0.07, 0.23) ng/mL and 0.99 (0.98, *>* 0.99) in the non-relapsing subgroup (*n* = 156), and 0.53 (0.39, 0.59) ng/mL and 0.95 (0.92, 0.98) in the relapsing subgroup (*n* = 10), respectively. Thus, the results from global fitting analysis demonstrate that the periodic dose model and the single dose model successfully recapitulate the observed patient-specific PSA dynamics. We also observe a good agreement between the fits provided by either model, as shown in Figure 2.

**Figure 2.**
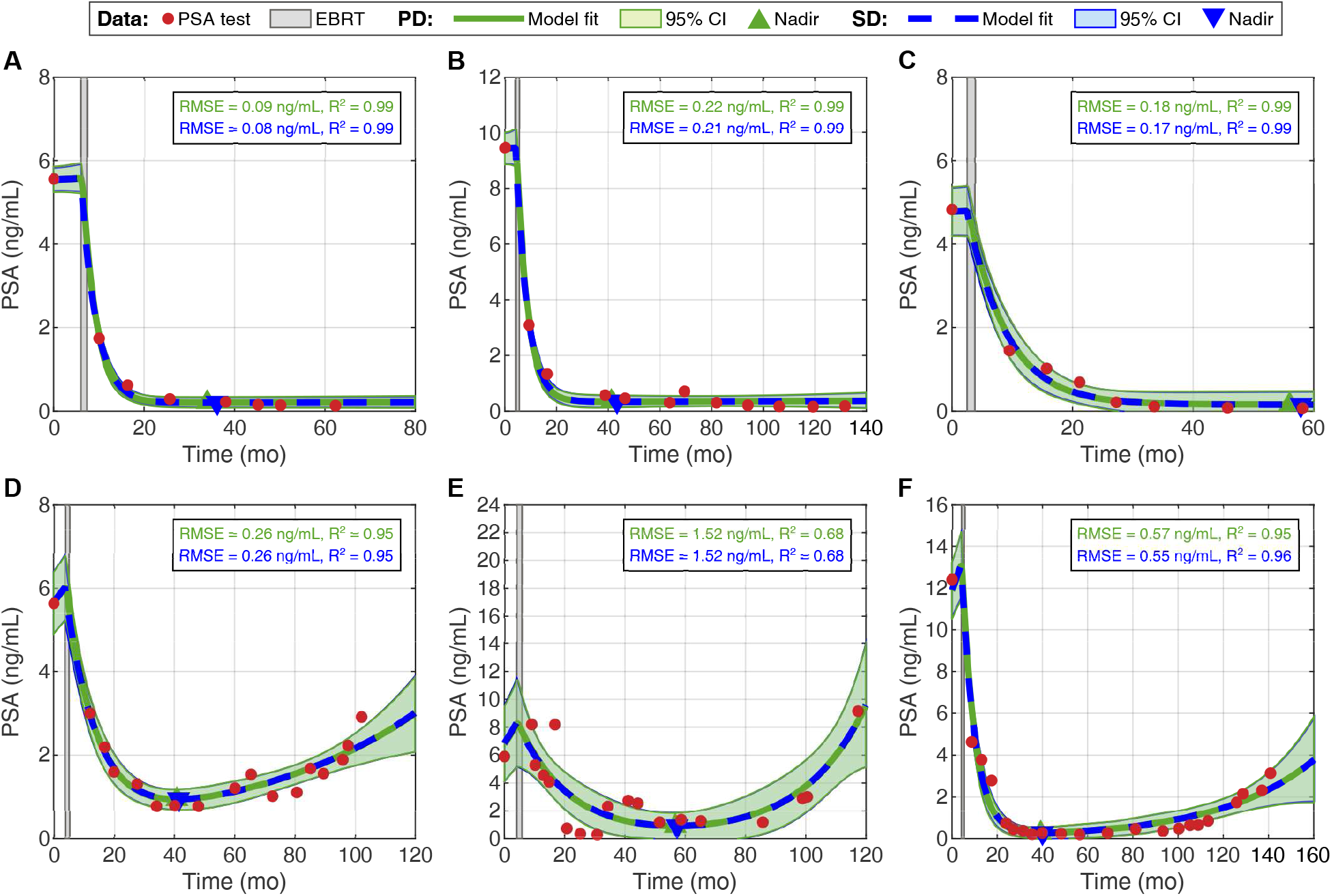
Mechanistic models reproduce the observed PSA dynamics during post-EBRT monitoring. Panels (A-C) show global fitting results for three non-relapsing patients, while panels (D-F) show similar results for three patients exhibiting a bio-chemical relapse. For each patient, the PSA measurements are plotted as red bullet points and EBRT is represented as a rectangular area shaded in light gray. The fits obtained with the periodic dose model (PD) are represented with a solid green line, while the fits produced by the single dose model (SD) are depicted as a dashed blue line. The same color scheme is also used for the 95% confidence intervals, which are depicted as the shaded areas around the model fits, and the font of the quality of fit metrics for either model. Additionally, each plot also includes the prediction of the patient’s PSA nadir using the periodic dose model (green upward triangle) and the single dose model (blue downward triangle). This figure illustrates the remarkable performance of our mechanistic models to fit patient-specific longitudinal PSA datasets collected during standard monitoring following EBRT. Additionally, there is a good agreement between both models, since corresponding fit curves and 95% confidence intervals practically coincide.

According to two-sided Wilcoxon rank-sum tests, the differences in quality of fit between the relapsing and the non-relapsing subgroups are significant for both the periodic dose model (RMSE: *p <* 0.001, *R*^2^: *p* = 0.0033) and the single dose model (RMSE: *p <* 0.001, *R*^2^: *p* = 0.0027). Corresponding one-sided tests show that superior fits are achieved for non-relapsing patients in terms of significantly lower RMSE and higher *R*^2^ with both the periodic dose model (RMSE: *p <* 0.001, *R*^2^: *p* = 0.0017) and the single dose model (RMSE: *p <* 0.001, *R*^2^: *p* = 0.0013). On a patient-wise basis, Wilcoxon signed-rank testing shows that the single dose model outperforms the periodic dose model in the non-relapsing subgroup, yielding significantly lower RMSE and higher *R*^2^ (*p <* 0.001 for both metrics in two- and one-sided tests). However, these results do not hold in the subgroup of relapsing patients. Additionally, no significant global differences in the RMSE and *R*^2^ distributions obtained using the periodic versus the single dose model are detected in any two-sided Wilcoxon rank-sum test performed across the whole cohort, the non-relapsing subgroup, and the relapsing one.

### Mechanistic models provide promising biomarkers of biochemical relapse

Following global fitting, we define a panel of model-based quantities of interest to examine their performance as biomarkers of biochemical relapse. As in [Lorenzo et al., 2019b], this panel is composed by three groups of quantities. First, we include the models’ parameters: the baseline PSA (*P*_0_), the proliferation rate of tumor cells (*ρ*_*s*_), the rate of EBRT-induced death of tumor cells (*ρ*_*d*_), and the fraction of tumor cells surviving to EBRT (*R*; *R* = *R*_*d*_ in the periodic dose model, *R* = *R*_*D*_ in the single dose model, and statistical analyses test 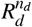 versus *R*_*D*_; see STAR Methods). Second, we also consider two non-dimensional metrics that represent the ratio or tumor cell proliferation to EBRT-induced death (*β*) and EBRT efficacy (*α*). Finally, we further use the models to calculate the PSA nadir (*P*_*n*_) and the time to PSA nadir since EBRT termination (Δ*t*_*n*_), which are two common metrics in the analysis of post-EBRT PSA dynamics [Lorenzo et al., 2019b; Freiberger et al., 2017; Zelefsky et al., 2005; Zumsteg et al., 2015; Ray et al., 2006; Bates et al., 2005; Cheung et al., 2006; Cavanaugh et al., 2004; Shi et al., 2013; Wein et al., 2012]. The interested reader is referred to the STAR Methods for further information on the definition of these model-based quantities of interest.

Figure 3 shows the boxplots of the distributions of all model-based quantities of interest across the cohort as well as the subgroups of non-relapsing and relapsing patients. Two-sided Wilcoxon rank-sum tests identify significant differences in the values obtained for *P*_0_, *ρ*_*s*_, *β*, and *P*_*n*_ using either the periodic dose model (*p* = 0.011, *<* 0.001, *<* 0.001, and *<* 0.001, respectively) or the single dose model (*p* = 0.0098, *<* 0.001, *<* 0.001, and *<* 0.001, respectively). Corresponding one-sided tests further show that relapsing patients exhibit higher *P*_0_, *ρ*_*s*_, *β*, and *P*_*n*_ than non-relapsing patients utilizing either the periodic dose model (*p* = 0.0053, *<* 0.001, *<* 0.001, and *<* 0.001, respectively) or the single dose model (*p* = 0.0049, *<* 0.001, *<* 0.001, and *<* 0.001, respectively). Additionally, a two-sided Wilcoxon rank-sum test identified significant differences in Δ*t*_*n*_ calculated with the single dose model (*p* = 0.040), but not in those obtained with the periodic dose model (*p* = 0.053). However, corresponding one-sided tests resulted in significantly lower values of Δ*t*_*n*_ for relapsing patients using either the periodic dose model (*p* = 0.027) or the single dose model (*p* = 0.020). As a result of this statistical analysis, we henceforth define *ρ*_*s*_, *β, P*_*n*_, and Δ*t*_*n*_ as model-based biomarkers of biochemical relapse. We do not consider *P*_0_ for two reasons. First, the baseline PSA measured in the clinic and reported in Table 1 (*P*_*d*_) was already significantly higher in relapsing patients (two-sided Wilcoxon rank-sum test *p* = 0.017). Additionally, *P*_0_ is bound to take values close to *P*_*d*_ as result of model fitting (see Figure 2 and [Lorenzo et al., 2019b]), thereby providing a limited contribution to the sequential patient-specific predictions of PSA dynamics and biochemical relapse obtained during the fitting-forecasting study.

**Figure 3.**
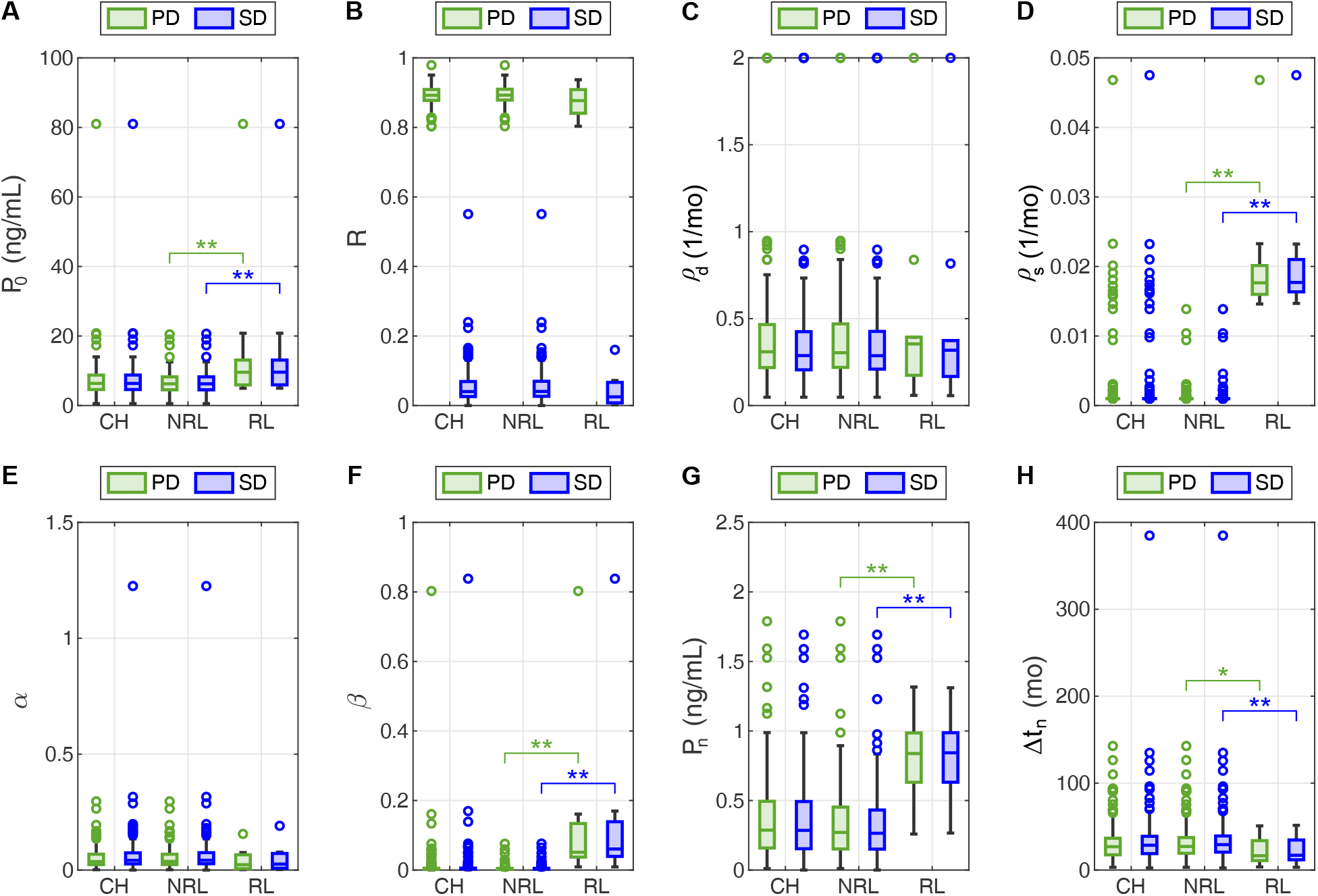
Distributions of the candidate model-based biomarkers of biochemical relapse obtained in the global fitting study. Each panel (A-H) shows the boxplots corresponding to the distribution of a model-based quantity of interest over the whole cohort (CH), the non-relapsing subgroup (NRL), and the relapsing one (RL). Green boxplots show results from the periodic dose model (PD), while blue boxplots correspond to the single dose model (SD). Outliers are represented as hollow circles. A double asterisk (**) indicates statistical significance in both two-sided and one-sided Wilcoxon rank-sum tests, while a single asterisk (*) denotes statistical significance only under a one-sided Wilcoxon rank-sum test (*p <* 0.05). (A) Initial PSA, *P*_0_. (B) Fraction of tumor cells surviving to EBRT (*R* = *R*_*d*_ in the PD model, *R* = *R*_*D*_ in the SD model, and here we test 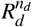 versus *R*_*D*_; see STAR Methods). (C) Rate of EBRT-induced death of tumor cells, *ρ*_*d*_. (D) Proliferation rate of tumor cells, *ρ*_*s*_. (E) Non-dimensional ratio *α*, representing EBRT efficiency (see STAR Methods). (F) Non-dimensional ratio *β*, representing the ratio of tumor cell proliferation to EBRT-induced death (see STAR Methods). (G) PSA nadir, *P*_*n*_. (H) Time to PSA nadir since EBRT termination, Δ*t*_*n*_. Table S1 further provides the median, interquartile range, and full range of the distribution of global fitting values for all the model-based quantities of interest.

Figure 4 shows the receiver operating characteristic (ROC) curves and corresponding optimal performance points for the model-based biomarkers identified from the analysis of the global fitting study results. For each biomarker and model, Table 2 further provides the area under the ROC curve (AUC) along with the optimal performance point threshold, sensitivity, and specificity. We observe that the ROC curves and optimal performance points obtained for each biomarker with either model are very similar, which suggests that the biomarker performance is independent of the model leveraged in its calculation. The shape of the ROC curves, the AUC, and the optimal performance points show that *ρ*_*s*_ exhibited the best performance in identifying relapsing patients, followed closely by *β*, then *P*_*n*_, and finally Δ*t*_*n*_, which only shows a mildly satisfactory performance. Notice that *ρ*_*s*_ operates as a perfect classifier, yielding maximal AUC along with 100% sensitivity and specificity at optimal performance point. This result can also be hinted in the boxplots of this biomarker shown in Figure 3, where a straight line could separate the values of *ρ*_*s*_ obtained with both models for relapsing and non-relapsing patients.

**Figure 4.**
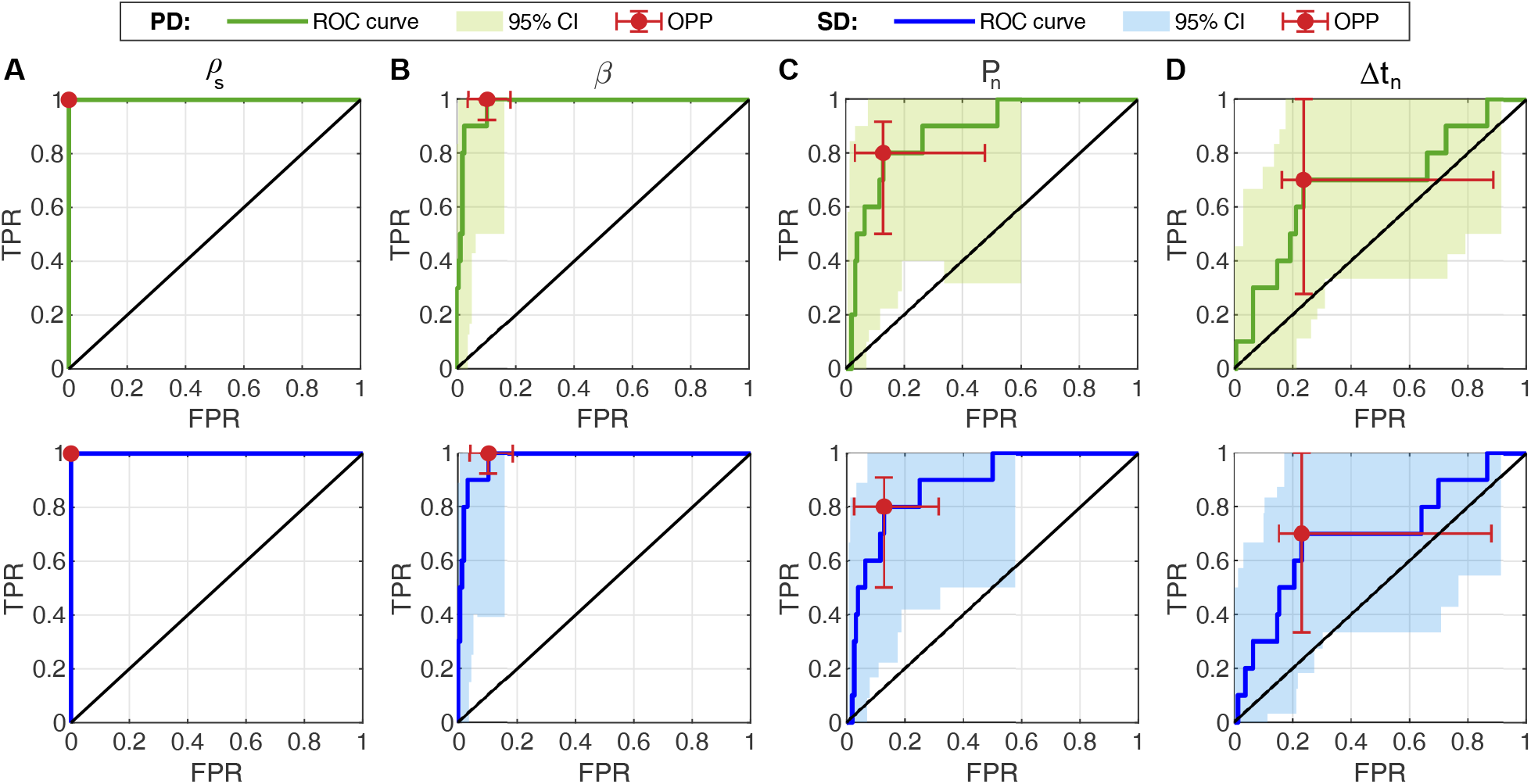
ROC curves for the model-based biomarkers of biochemical relapse identified in the analysis of the global fitting study results. Panels (A-D) show the ROC curves and 95% bootstrap confidence intervals (CI) for each biomarker constructed using the global fitting results from the periodic dose model (PD, top row, in green) single dose model (SD, bottom row, in blue). In each plot, the vertical axis quantifies the true positive rate (TPR, i.e., sensitivity), while the horizontal axis measures the false positive rate (FPR, i.e., 1-specificity). The solid black line represents the unity line. The optimal performance point (OPP) and corresponding 95% bootstrap confidence intervals along both axes are depicted as a red solid point and errorbars, respectively. (A) Proliferation rate of tumor cells, *ρ*_*s*_. (B) Non-dimensional ratio *β*, representing the ratio of tumor cell proliferation to EBRT-inuced death (see STAR Methods). (C) PSA nadir, *P*_*n*_. (D) Time to PSA nadir since EBRT termination, Δ*t*_*n*_.

**Table 2.** Analysis of the ROC curves of the model-based biomarkers of biochemical relapse identified in the global fitting study. Values in brackets are 95% bootstrap confidence intervals of the reported metrics. AUC: area under the ROC curve.

| Metric | Biomarker |  |  |  |
| --- | --- | --- | --- | --- |
| | $\rho_s$ | $\beta$ | $P_n$ | $\Delta t_n$ |
| **Periodic dose model** |  |  |  |  |
| AUC | 1.00 [1.00, 1.00] | 0.98 [0.93, 1.00] | 0.88 [0.68, 0.95] | 0.68 [0.45, 0.85] |
| Optimal performance point |  |  |  |  |
| Sensitivity | 1.00 [1.00, 1.00] | 1.00 [0.92, 1.00] | 0.80 [0.50, 0.92] | 0.70 [0.28, 1.00] |
| Specificity | 1.00 [1.00, 1.00] | 0.90 [0.82, 0.96] | 0.87 [0.52, 0.97] | 0.76 [0.11, 0.84] |
| Value | 14.6 [13.9, 14.6] $\cdot 10^{-3}$/mo | 9.0 [6.7, 22.8] $\cdot 10^{-3}$ | 0.6 [0.3, 0.9] ng/mL | 19.0 [15.9, 53.6] mo |
| **Single dose model** |  |  |  |  |
| AUC | 1.00 [1.00, 1.00] | 0.98 [0.94, 1.00] | 0.88 [0.71, 0.95] | 0.69 [0.47, 0.87] |
| Optimal performance point |  |  |  |  |
| Sensitivity | 1.00 [1.00, 1.00] | 1.00 [0.92, 1.00] | 0.80 [0.50, 0.91] | 0.70 [0.33, 1.00] |
| Specificity | 1.00 [1.00, 1.00] | 0.90 [0.82, 0.96] | 0.87 [0.68, 0.97] | 0.77 [0.12, 0.85] |
| Value | 14.7 [13.9, 14.7] $\cdot 10^{-3}$/mo | 9.0 [7.6, 23.4] $\cdot 10^{-3}$ | 0.6 [0.4, 0.9] ng/mL | 19.8 [16.2, 54.7] mo |

### Mechanistic models accurately forecast short-term PSA dynamics after EBRT

To assess the predictive potential of the model-based biomarkers of biochemical relapse identified in the global fitting study, we first analyze the accuracy of the predictions of PSA dynamics obtained with our mechanistic models. To this end, we run a fitting-forecasting study as follows: first, we estimate the models parameters by fitting them to a subset of the earliest PSA values for each patient, we then calculate the corresponding personalized models’ forecasts of PSA dynamics, and we finally compare each of these predictions to the remainder of the patient’s PSA data in posterior dates. For each patient, we perform this calculation in a collection of sequential scenarios: we start using a minimum of five PSA values for fitting (*n*_*P,fit*_ = 5) including the baseline PSA at diagnosis (*P*_*d*_), and we progressively increase *n*_*P,fit*_ until only one PSA value is left for assessing the models’ predictions (i.e., *n*_*P,fit*_ = 5, …, *n*_*P*_ − 1 for each patient; see STAR Methods for further methodological details). Additionally, in this study we consider a maximum *n*_*P,fit*_ = 21, since this is the last scenario for which there is at least one relapsing and one non-relapsing patient with at least one remaining PSA value to assess the models’ forecasts.

Figure 5 illustrates representative results from the fitting-forecasting study for the relapsing and nonrelapsing patients considered in Figure 2 in the global fitting study. The results in Figure 5 show that our models can accurately predict PSA in the short term after the date of the last PSA used in model fitting. This is the case for both non-relapsing and relapsing patients across the different *n*_*P,fit*_ scenarios considered in our analysis. While the prediction of the long-term PSA plateau in the non-relapsing subgroup is accurate even with a low *n*_*P,fit*_ (see Figure 5(A-C)), our models may require exposition to an incipient rising trend to accurately identify a biochemical relapse and estimate long-term PSA values (see Figure 5(D-F) versus Figure 5(G-I)). However, Figure 5(G-I) further shows that our model forecasts are able to identify rising PSA dynamics in relapsing patients earlier than standard clinical detection methods (e.g., using the nadir+2 ng/mL criterion). Additionally, as in the global fitting study, we observe an overall good agreement between the fitting and forecasting results obtained with the periodic dose and the single dose models. Comparing Figure 5 and Figure 2, we also observe that, as the number of PSA values used for model fitting increases (i.e., for higher *n*_*P,fit*_), the uncertainty in the model predictions of PSA decreases accordingly and progressively approaches the level of uncertainty obtained in the global fitting scenario.

**Figure 5.**
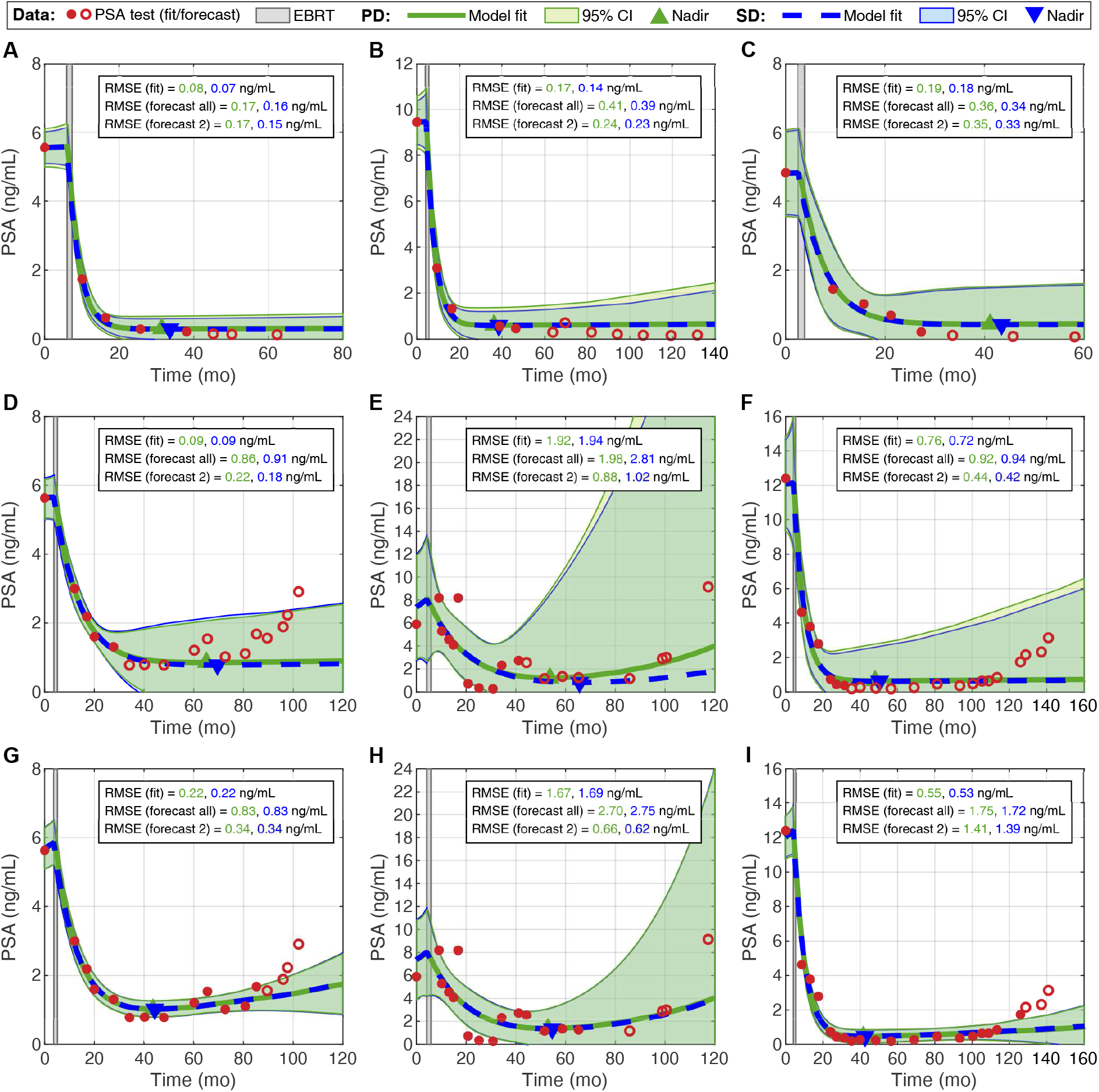
Mechanistic models provide personalized forecasts of PSA dynamics during post-EBRT monitoring. Panels (A-C) and (D-F) show an early forecast of the PSA dynamics obtained with our mechanistic models for the same non-relapsing and relapsing patients considered in Figure 2, respectively. The early model forecasts for the non-relapsing patients accurately predict both short and long-term PSA dynamics. However, our early forecasts of PSA are only accurate in the short term for the relapsing patients. Panels (G-I) show the model forecasts of PSA dynamics obtained at a later date in the course of post-EBRT monitoring for the same non-relapsing patients (i.e., in a higher *n*_*P,fit*_ scenario). In this case, while the models can still predict only the following short-term PSA values accurately, they can already detect a rising PSA trend earlier than standard clinical practice (e.g., using the nadir+2 ng/mL criterion). Moreover, the PSA predictions and corresponding 95% confidence intervals in this figure demonstrate an overall good agreement between the models in forecasting PSA dynamics. For each patient, the PSA measurements used for model fitting are plotted as red bullet points, while the remainder PSA data to assess model predictions are represented as hollow red circles. EBRT is represented as a light gray rectangular area. The fits and forecasts obtained with the periodic dose model (PD) are represented with a solid green line, while the corresponding results produced by the single dose model (SD) are depicted with a dashed blue line. The same color scheme is also used for the 95% confidence intervals, which are depicted as shaded areas around the model fits, and the font of the fitting and forecasting RMSEs for either model. The forecasting RMSE is reported for both all the remaining PSA values and the immediately next two PSA values after the date of the last PSA used for model fitting. Additionally, each plot also includes the prediction of the patient’s PSA nadir using the periodic dose model (green upward triangle) and the single dose model (blue downward triangle).

To analyze the distribution of the global RMSE values across all the fitting and forecasting scenarios, we pool the corresponding results obtained with either model for all *n*_*P,fit*_ cases run for each patient in the cohort. Since our mechanistic models provide an accurate prediction of short-term dynamics in both non-relapsing and relapsing patients, we will focus the analysis of the forecasting results on the subsequent two PSA values that were not used in model fitting in each *n*_*P,fit*_ scenario for each patient. Hence, this two-value forecast corresponds to a short-term prediction of PSA dynamics over approximately a one year horizon according to the PSA test frequency in our cohort (see Table 1). Note that not all patients for which it is possible to fit *n*_*P,fit*_ PSA values are eligible for a short-term prediction of two PSA values, because of limited PSA data for forecast validation. For the sake of completeness, Figure S1 further depicts the boxplots of the RMSE for model fitting and short-term PSA prediction in each fitting-forecasting scenario across the whole cohort (*n*_*P,fit*_ = 5, …, 21). Additionally, Tables S2 and S3 summarize these RMSE distributions for the whole cohort as well as the non-relapsing and relapsing subgroups.

Using the periodic dose model, the median and interquartile range of the RMSE of the model fits are 0.16 (0.08, 0.25) ng/mL across the whole cohort (*n* = 1043), 0.15 (0.08, 0.23) ng/mL in the non-relapsing subgroup (*n* = 949), and 0.34 (0.16, 0.76) ng/mL in the relapsing subgroup (*n* = 94). For the short-term prediction of PSA using the patient-specific periodic dose model fits, the corresponding median and interquartile range of the RMSE are 0.18 (0.08, 0.36) ng/mL across the whole cohort (*n* = 878), 0.17 (0.07, 0.32) ng/mL in the non-relapsing subgroup (*n* = 794), and 0.67 (0.22, 1.20) ng/mL in the relapsing subgroup (*n* = 84). Furthermore, the median and interquartile range of the RMSE of the single dose model fits are 0.15 (0.08, 0.25) ng/mL across the whole cohort (*n* = 1043), 0.14 (0.07, 0.22) ng/mL in the non-relapsing sub-group (*n* = 949), and 0.34 (0.17, 0.72) ng/mL in the relapsing subgroup (*n* = 94). Using the patient-specific single dose model fits to perform a short-term forecast of PSA, the corresponding median and interquartile range of the RMSE are 0.18 (0.08, 0.34) ng/mL across the whole cohort (*n* = 878), 0.16 (0.07, 0.31) ng/mL in the non-relapsing subgroup (*n* = 794), and 0.63 (0.22, 1.19) ng/mL in the relapsing subgroup (*n* = 84). These RMSE results demonstrate that our models can reproduce the observed PSA dynamics, and that the resulting personalized models can yield a reasonably accurate prediction of PSA to inform the monitoring strategy for each patient (see Discussion).

Considering the pooled results across all *n*_*P,fit*_ cases, the RMSE values of the model fits and the short-term PSA predictions obtained for non-relapsing patients were significantly lower than those obtained for relapsing patients. This happens using both the periodic dose and the single dose model (*p <* 0.001 in all corresponding two- and one-sided Wilcoxon rank-sum tests). On a patient-wise basis, Wilcoxon signed-rank testing shows that the single dose model rendered significantly lower RMSE values than the periodic dose model during both fitting and short-term forecasting in the non-relapsing subgroup (*p <* 0.001 for both two- and one-sided tests). However, this result does not hold in the relapsing subgroup. Furthermore, two-sided Wilcoxon rank-sum tests do not detect significant differences between the distributions of the global RMSE values of the model fits obtained with either model across the whole cohort, the non-relapsing subgroup, and the relapsing subgroup. The same result is also obtained when comparing the short-term PSA forecasts obtained with either model. Using two-sided Wilcoxon rank-sum test to compare the global RMSE distributions obtained for model fits against those calculated for the short-term PSA predictions, we obtain significant differences across the whole cohort, the non-relapsing subgroup, and the relapsing subgroup for both the periodic dose model (*p <* 0.001, *p <* 0.001, and *p* = 0.018, respectively) and the single dose model (*p <* 0.001, *p <* 0.001, and *p* = 0.025, respectively). Corresponding one-sided tests further confirm that the RMSE values for the model fits are significantly lower than those of the short-term PSA predictions for both the periodic dose model (*p <* 0.001, *p <* 0.001, and *p* = 0.0092, respectively) and the single dose model (*p <* 0.001, *p <* 0.001, and *p* = 0.013, respectively).

### Early estimates of model-based biomarkers accurately predict biochemical relapse earlier than standard practice

The patient-specific model fits obtained for each *n*_*P,fit*_ scenario in the fitting-forecasting study provide a set of values for the model-based biomarkers of biochemical relapse identified during global fitting (i.e., *ρ*_*s*_, *β, P*_*n*_, and Δ*t*_*n*_). Since each *n*_*P,fit*_ scenario can only be performed once the latest PSA value used for model fitting is collected, the corresponding estimates of the biomarkers are associated with its collection date. Thus, we proceed to analyze whether early estimates of our model-based biomarkers enable an accurate identification of relapsing patients and whether they anticipate the detection of relapse with respect to standard clinical practice. To further motivate this analysis, we recall that, for the three relapsing patients shown in Figure 5(G-I), our models predict a rising PSA trend before the usual nadir+2 ng/mL criterion is satisfied.

To assess the performance of our early model-based biomarker estimates as biochemical relapse classifiers, we perform a ROC curve analysis. We first pool all the values obtained for each biomarker in all the *n*_*P,fit*_ scenarios across all patients. Then, we construct the ROC curve by using each pooled biomarker value as a threshold that we compare to the corresponding *n*_*P,fit*_ values obtained for each patient in the fitting-forecasting study (*n*_*P,fit*_ = 5, …, *n*_*P*_ − 1). If any of these patient-specific *n*_*P,fit*_ values satisfies the classification criterion for the biomarker (i.e., a larger *ρ*_*s*_, *β*, and *P*_*n*_ or a lower Δ*t*_*n*_), then we identify the patient as relapsing under the considered threshold. As in the global fitting study, we perform this ROC curve analysis for each model separately. We refer the interested reader to the STAR Methods for further methodological details.

Figure 6 shows the ROC curve and optimal performance point for each biomarker obtained by using both the periodic and the single dose model results of the fitting-forecasting study. Table 3 further reports the corresponding AUC value along with the optimal performance point threshold, sensitivity, and specificity. Additionally, both Figure 6 and Table 3 represent the ability of the optimal performance point threshold identified in the global fitting study to classify biochemical relapse using the early biomarker estimates from the fitting-forecasting study. The ROC curves and optimal performance points obtained with either model are virtually equivalent, as we had previously observed in the ROC curve analysis of the global fitting results (see Figure 4 and Table 2). Furthermore, we obtain again that the best performing biomarkers are *ρ*_*s*_ and *β*, followed by *P*_*n*_, and finally Δ*t*_*n*_, which exhibits a comparably poorer classifying ability with respect to the other three biomarkers. Comparing the ROC curve metrics obtained in the global fitting study and the fitting-forecasting study, the AUC is slightly lower when the biomarkers obtained with either model are assessed to early identify biochemical relapse. While parameter *ρ*_*s*_ acted as a perfect classifier in the global fitting study, we observe a minor loss of specificity when it is leveraged as an early biomarker for biochemical relapse. Note also that the optimal threshold required to provide an early classification of biochemical relapse with *ρ*_*s*_ is notably lower with respect to the one obtained in the global fitting study. Thus, a less conservative threshold is needed to provide an early identification of biochemical relapse using *ρ*_*s*_. This is also suggested by comparing the model predictions for the relapsing patients in Figure 2(D-F) and Figure 5(G-I). Conversely, the *β* ratio exhibits a slightly better performance in the fitting-forecasting study due to a moderate increase in specificity, and the optimal threshold stays in the same order of magnitude. This last observation also holds for the PSA nadir *P*_*n*_, which shows maximal sensitivity in the fitting-forecasting study. However its specificity to early detect biochemical relapse is lower than in the global fitting study. Finally, Δ*t*_*n*_ also exhibits a lower specificity in the fitting-forecasting study than in the global fitting scenario.

**Figure 6.**
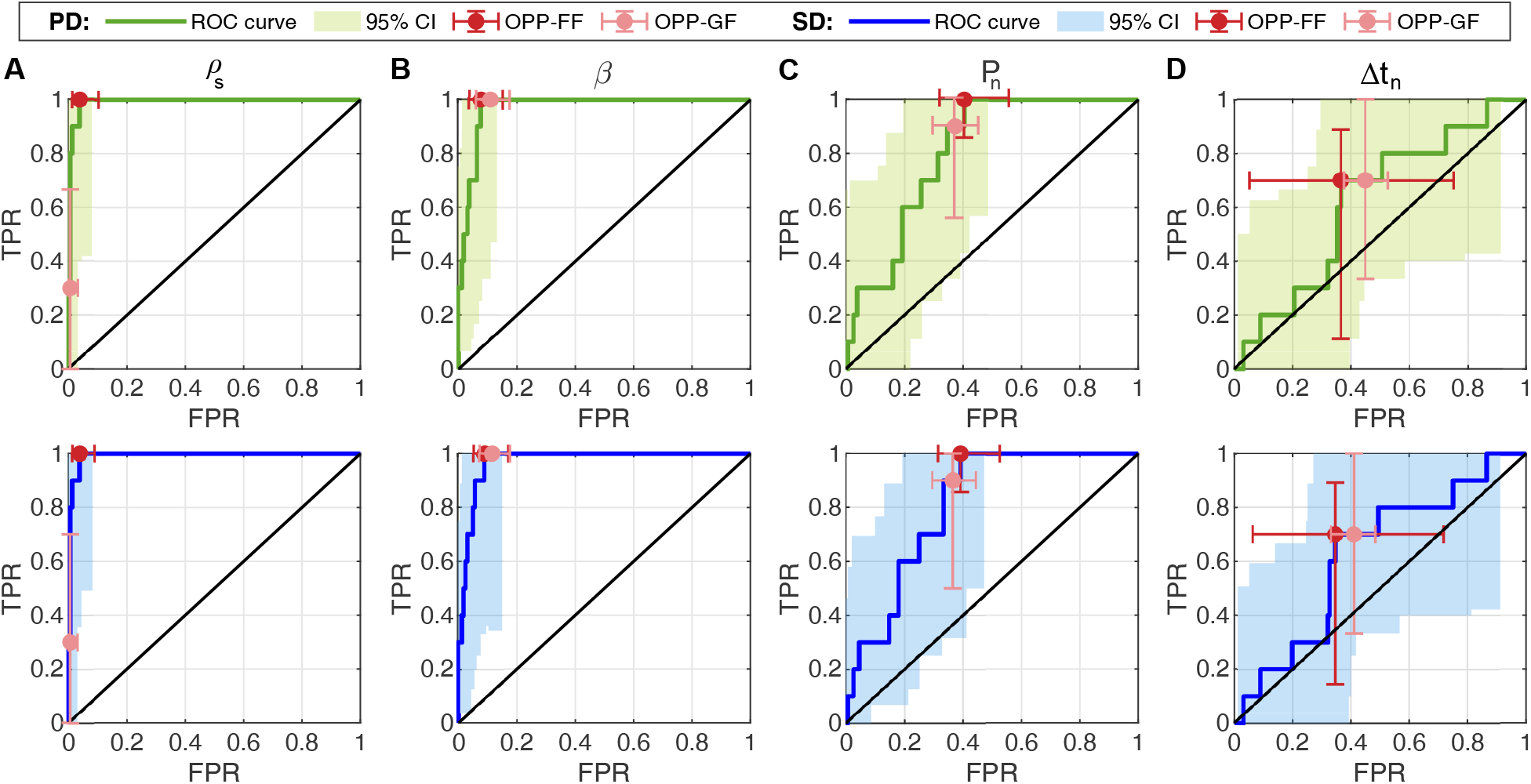
ROC curves for the early estimates of the biochemical relapse biomarkers obtained in the fitting-forecasting studies. Panels (A-D) show the ROC curves and 95% bootstrap confidence intervals (CI) for each biomarker constructed using the fitting-forecasting results from the periodic dose model (PD, top row, in green) single dose model (SD, bottom row, in blue). In each plot, the vertical axis quantifies the true positive rate (TPR, i.e., sensitivity), while the horizontal axis measures the false positive rate (FPR, i.e., 1-specificity). The solid black line represents the unity line. The optimal performance point (OPP-FF) and corresponding 95% bootstrap confidence intervals along both axes are depicted as a red solid point and errorbars, respectively. Additionally, the optimal performance point identified during global fitting (OPP-GF) and corresponding 95% bootstrap confidence intervals are represented with a pink solid point and errorbars, respectively. (A) Proliferation rate of tumor cells, *ρ*_*s*_. (B) Non-dimensional ratio *β*, representing the ratio of tumor cell proliferation to EBRT-inuced death (see STAR Methods). (C) PSA nadir, *P*_*n*_. (D) Time to PSA nadir since EBRT termination, Δ*t*_*n*_.

**Table 3.** Analysis of the ROC curves of the model-based biomarkers to identify early biochemical relapse using the fitting-forecasting study results. Values in brackets are 95% bootstrap confidence intervals of the reported metrics. For each biomarker, the global fitting threshold was fixed to the optimal cutoff reported in Table 2 in order to determine the corresponding 95% bootstrap confidence intervals for its sensitivity and specificity using the fitting-forecasting study results. AUC: area under the ROC curve.

| Metric | Biomarker |  |  |  |
| --- | --- | --- | --- | --- |
| | $\rho_s$ | $\beta$ | $P_n$ | $\Delta t_n$ |
| **Periodic dose model** |  |  |  |  |
| AUC | 0.99 [0.97, 1.00] | 0.97 [0.93, 0.99] | 0.81 [0.71, 0.90] | 0.62 [0.42, 0.77] |
| Optimal performance point |  |  |  |  |
| *Fitting-forecasting study* |  |  |  |  |
| Sensitivity | 1.00 [1.00, 1.00] | 1.00 [1.00, 1.00] | 1.00 [0.86, 1.00] | 0.70 [0.11, 0.89] |
| Specificity | 0.96 [0.90, 0.99] | 0.92 [0.85, 0.96] | 0.60 [0.45, 0.68] | 0.63 [0.25, 0.95] |
| Value | $5.6 [2.0, 9.1] \cdot 10^{-3}/\text{mo}$ | $13.0 [8.8, 16.4] \cdot 10^{-3}$ | 0.6 [0.5, 0.7] ng/mL | 16.7 [8.3, 36.6] mo |
| *Global fitting study* |  |  |  |  |
| Sensitivity | 0.30 [0.00, 0.67] | 1.00 [1.00, 1.00] | 0.90 [0.57, 1.00] | 0.70 [0.33, 1.00] |
| Specificity | 0.99 [0.97, 1.00] | 0.89 [0.83, 0.94] | 0.63 [0.55, 0.70] | 0.55 [0.47, 0.62] |
| Value | $14.6 \cdot 10^{-3} \text{ 1/mo}$ | $9.0 \cdot 10^{-3}$ | 0.6 ng/mL | 19.0 mo |
| **Single dose model** |  |  |  |  |
| AUC | 0.99 [0.97, 1.00] | 0.97 [0.93, 0.99] | 0.81 [0.70, 0.90] | 0.63 [0.44, 0.78] |
| Optimal performance point |  |  |  |  |
| *Fitting-forecasting study* |  |  |  |  |
| Sensitivity | 1.00 [1.00, 1.00] | 1.00 [1.00, 1.00] | 1.00 [0.86, 1.00] | 0.70 [0.14, 0.89] |
| Specificity | 0.96 [0.91, 0.99] | 0.91 [0.83, 0.95] | 0.61 [0.47, 0.69] | 0.65 [0.28, 0.94] |
| Value | $5.7 [2.4, 10.0] \cdot 10^{-3}/\text{mo}$ | $13.3 [7.6, 17.4] \cdot 10^{-3}$ | 0.6 [0.6, 0.7] ng/mL | 18.1 [7.9, 39.6] mo |
| *Global fitting study* |  |  |  |  |
| Sensitivity | 0.30 [0.00, 0.70] | 1.00 [1.00, 1.00] | 0.90 [0.50, 1.00] | 0.70 [0.33, 1.00] |
| Specificity | 0.99 [0.97, 1.00] | 0.88 [0.82, 0.93] | 0.63 [0.56, 0.71] | 0.59 [0.52, 0.67] |
| Value | $14.7 \cdot 10^{-3} \text{ 1/mo}$ | $9.0 \cdot 10^{-3}$ | 0.6 ng/mL | 19.8 mo |

We further leverage the optimal threshold calculated for the ROC curve of each model-based biomarker in the fitting-forecasting study to assess whether it may enable an earlier detection of biochemical relapse than standard clinical criteria (e.g., nadir+2 ng/mL). To this end, we define a metric termed *Days Gained to Biochemical Relapse Diagnosis* (DGBRD) for each biomarker and for every patient in the relapsing subgroup. This metric is computed as the difference between the reported date of biochemical relapse and the earliest date in which each biomarker classifies a patient as relapsing according to the optimal threshold calculated in the ROC curve analysis of the fitting-forecasting study. A positive value of DGBRD indicates that a biomarker enables the early detection of biochemical relapse with respect to standard clinical practice. Note that the second date in the DGBRD definition corresponds to one of the dates in which PSA was measured for each relapsing patient, since these are the dates at which we calculate the model fits in each of the *n*_*P,fit*_ scenarios of the fitting-forecasting study.

Figure 7 shows the distribution of the DGBRD for each biomarker calculated with both the periodic and the single dose model, and Table 4 reports the corresponding median, interquartile range, and full range. Using one-sided signed-rank Wilcoxon tests on median larger than zero, we obtain that the early estimates of *ρ*_*s*_, *β*, and *P*_*n*_ provide a significantly earlier detection of biochemical relapse than standard clinical practice either leveraging the periodic dose model (*p* = 0.0029, 0.0029, and 0.0056, respectively) or the single dose model (*p* = 0.0029, 0.0029, and 0.0098, respectively) for their calculation. For both models, we observe that, while *ρ*_*s*_ and *β* exhibited the best classifier performance in the ROC curve analysis (see Figure 6 and Table 3), *P*_*n*_ is the biomarker yielding the earliest identification of biochemical relapse. Indeed, two-sided Wilcoxon signed-rank tests identify significant patient-wise differences in the DGBRD for *P*_*n*_ with respect to those calculated for *ρ*_*s*_, *β*, and Δ*t*_*n*_ for both the periodic dose model (*p* = 0.016, 0.016, and 0.0078, respectively) and the single dose model (*p* = 0.019, 0.019, and 0.016, respectively). Corresponding one-sided tests confirm that the DGBRD calculated for *P*_*n*_ provided a significantly earlier identification of biochemical relapse than the DGBRD obtained for *ρ*_*s*_, *β*, and Δ*t*_*n*_ for each patient with both the periodic dose model (*p* = 0.0078, 0.0078, and 0.0039, respectively) and the single dose model (*p* = 0.0098, 0.0098, and 0.0078, respectively). The comparison of the *DGBRD* for *ρ*_*s*_ and *β* with respect to the DGBRD for Δ*t*_*n*_ from the single dose model was not significant under two-sided Wilcoxon signed-rank testing. However, the corresponding one-sided tests do identify a significantly larger DGBRD for *ρ*_*s*_ and *β* (*p* = 0.039 and *p* = 0.027, respectively). For the periodic dose model, this observation only holds for the comparison of the DGBRD values of *β* and Δ*t*_*n*_ (*p* = 0.027 for the one-sided Wilcoxon signed-rank test). Comparing the overall DGBRD distributions between the biomarkers under two-sided Wilcoxon rank-sum testing only the DGBRD values of *P*_*n*_ and Δ*t*_*n*_ show significant differences for the periodic dose model (*p* = 0.022). The corresponding one-sided tests identify a significantly larger DGBRD for *P*_*n*_ than for Δ*t*_*n*_ for both models (*p* = 0.011 and *p* = 0.026, respectively). In addition, no significant differences are identified between the DGBRD values obtained for each biomarker with both models by using either two-sided Wilcoxon rank-sum or signed-rank tests.

**Figure 7.**
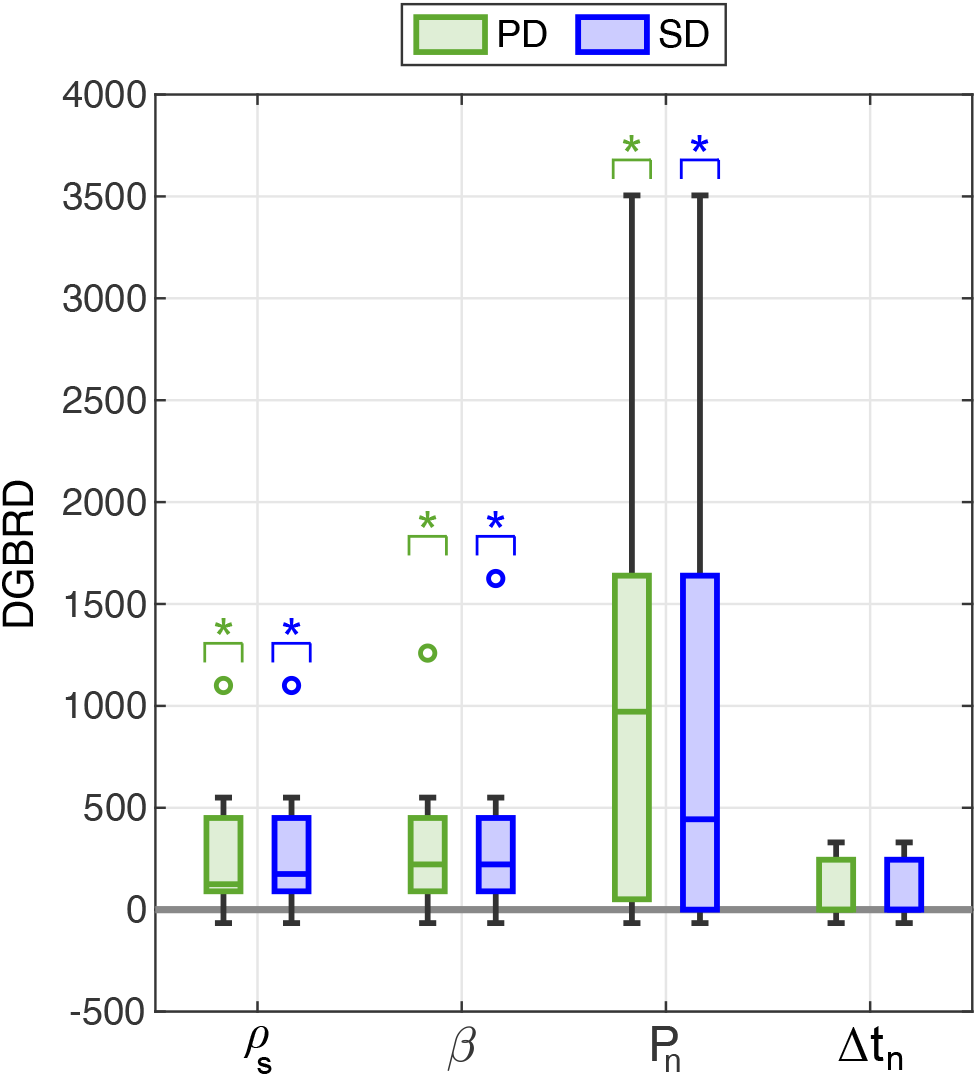
Distribution of the Days Gained to Biochemical Relapse Diagnosis (*DGBRD*) for the model-based biomarkers of biochemical relapse. The boxplots in this figure represent the distribution of the *DGBRD* in the relapsing subgroup (*n* = 10) obtained for each biomarker of biochemical relapse by leveraging the optimal threshold calculated in the ROC curve analysis of the fitting-forecasting study results for both the periodic dose and the single dose model. From left to right: proliferation rate of tumor cells *ρ*_*s*_, non-dimensional ratio *β* (i.e., ratio of tumor cell proliferation to EBRT-induced death; see STAR Methods), PSA nadir *P*_*n*_, and time to PSA nadir since EBRT termination Δ*t*_*n*_. Green boxplots show results from the periodic dose model (PD), while blue boxplots correspond to the single dose model (SD). Outliers are represented as hollow circles. A single asterisk (*) denotes statistical significance in one-sided Wilcoxon signed-rank tests for a median larger than zero (*p <* 0.05).

**Table 4.** Distribution of the Days Gained to Biochemical Relapse Diagnosis (*DGBRD*) in the relapsing subgroup (*n* = 10) for each biomarker obtained leveraging the optimal threshold calculated in the ROC curve analysis of the fitting-forecasting results for both the periodic dose and the single dose model. IQR: interquartile range.

| Biomarker | Periodic dose model |  |  | Single dose model |  |  |
| --- | --- | --- | --- | --- | --- | --- |
|  | Median | IQR | Range | Median | IQR | Range |
| $\rho_s$ | 125 | (90, 450) | (-66, 1100) | 175 | (90, 450) | (-66, 1100) |
| $\beta$ | 222 | (90, 450) | (-66, 1259) | 222 | (90, 450) | (-66, 1625) |
| $P_n$ | 971.5 | (51, 1639) | (-66, 3505) | 443.5 | (0, 1639) | (-66, 3505) |
| $\Delta t_n$ | 0 | (0, 245) | (-66, 330) | 0 | (0, 245) | (-66, 330) |

## Discussion

### Patient-specific predictions of PSA based on mechanistic models to design personalized PSA monitoring strategies

The detection of a consistent rising trend in PSA constitutes a biochemical relapse, which is indicative of a potential PCa recurrence [Wein et al., 2012; Mottet et al., 2021; Cornford et al., 2021]. Here, we posit that patient-specific forecasts of PSA evolution obtained *via* mechanistic modeling of post-EBRT PSA dynamics can accurately identify relapsing patients earlier than the current PSA threshold criteria for detection (e.g., nadir+2 ng/mL) [Wein et al., 2012; Roach et al., 2006; Cornford et al., 2021]. To this end, we leverage our previously proposed mechanistic models of PSA dynamics after EBRT [Lorenzo et al., 2019b]. These models feature key advantages for their clinical use to forecast PSA during post-EBRT patient monitoring. First, they are defined upon the essential mechanisms underlying the tumor response to radiation and the ensuing changes in PSA dynamics after EBRT conclusion (see STAR Methods and Figure 1). Consequently, our models have a simple formulation with only four parameters to be identified patient-wise from the longitudinal PSA data that are routinely collected during standard patient follow-up (see STAR Methods and Figure 1). Finally, the mathematical solutions to our model span the plateauing trend observed in non-relapsing patients as well as the biexponential response expected for relapsing patients [Zagars and Pollack, 1997; Cox et al., 1994; Hanlon et al., 1998; Vollmer and Montana, 1999; Taylor et al., 2005].

In this work, we first demonstrate that our models exhibit a remarkable accuracy in reproducing the complete post-EBRT PSA dynamics observed in a new patient cohort from a different center to the one from our previous study [Lorenzo et al., 2019b], thereby providing preliminary evidence of cross-institutional validation. We further demonstrate that our models can provide reasonably accurate short-term forecasts of PSA for both non-relapsing and relapsing patients over the course of post-EBRT monitoring. To this end, we focused on predicting the immediately next two PSA values that were not used to parameterize our models in the serial *n*_*P,fit*_ scenarios run for each patient (*n*_*P,fit*_ = 5, …, *n*_*P*−1_) during the fitting-forecasting study. As noted in the Results section, our short-term PSA predictions correspond to a time horizon of approximately one year according to the PSA testing frequency in our cohort (see Table 1). This prediction time horizon overlaps with standard PSA monitoring protocols, which usually define routine PSA tests more frequently over the first years following EBRT termination (e.g., every 3 to 6 months) and more sparsely afterwards (e.g., every 6 to 12 months) [Hamdy et al., 2016; Wein et al., 2012], unless the collected PSA values rise suspicion of a potential relapse that would warrant more frequent testing (e.g., moderately high PSA levels, an incipient rising trend) [Freiberger et al., 2017; Zelefsky et al., 2005; Zumsteg et al., 2015; Ray et al., 2006; Bates et al., 2005; Cheung et al., 2006; Cavanaugh et al., 2004; Shi et al., 2013; Wein et al., 2012]. Thus, our model forecasts could facilitate the systematic design of personalized monitoring plans, which can be adapted as the collection of further PSA data become available to inform our models. For instance, the consistent prediction of a plateauing trend during post-EBRT monitoring could be used to extend the time interval between consecutive PSA tests. Conversely, the detection of rising PSA dynamics would motivate a decision to prescribe more frequent PSA tests to confirm a biochemical relapse and then proceed to assess PCa recurrence (e.g., *via* biopsy and medical imaging) [Cornford et al., 2021; Wein et al., 2012]. Hence, our mechanistic modeling forecasts could advance patient monitoring after EBRT from routine and observational PSA testing to a dynamic predictive paradigm optimizing PSA data collection on a patient-specific basis.

### Promising model-based biomarkers to early detect biochemical relapse

In addition to the explicit forecast of PSA values with our models, we further explore the performance of model-based biomarkers of biochemical relapse. By analyzing our global model fits to the whole PSA dataset available for each patient, we identify four candidate biomarkers of biochemical relapse: the proliferation rate of tumor cells (*ρ*_*s*_), its ratio to the radiation-induced tumor cell death rate (*β*), the PSA nadir (*P*_*n*_), and the time to PSA nadir since EBRT termination (Δ*t*_*n*_).

As in [Lorenzo et al., 2019b], we observe a superior performance in classifying relapsing patients for the biomarkers that are directly related to the underlying tumor dynamics (*ρ*_*s*_ and *β*) than for the biomarkers that are more closely linked to PSA dynamics (*P*_*n*_ and Δ*t*_*n*_; see Figure 4 and Table 2). In particular, our results show that relapsing patients exhibit high *ρ*_*s*_ and *β* (see Figure 3 and Table S1). A high proliferation activity measured *via* Ki-67 staining in PCa tissue samples has been correlated with worse prognosis, radiotherapeutic outcome, and survival [Cowen et al., 2002; Li et al., 2004; Berlin et al., 2017]. An elevated tumor cell proliferation has also been correlated with an increased risk of PCa aggressiveness in terms of a higher Gleason score [Tretiakova et al., 2016], which is a histopathological metric that is ubiquitously used in the clinical management of PCa. In particular, a higher pre-treatment Gleason score has been linked to higher probability of both local and distant PCa recurrence [Zumsteg et al., 2015; Zelefsky et al., 2005; Wein et al., 2012]. The measurement of both the Ki-67 staining index and the Gleason score require an invasive approach to extract patient-specific tissue samples. However, our mechanistic models could provide a non-invasive surrogate method to estimate the proliferation activity of PCa in terms of *ρ*_*s*_ and *β*, and thereby refine the estimation of patient-specific prognosis to guide therapeutic planning [Cowen et al., 2002; Li et al., 2004; Berlin et al., 2017].

*P*_*n*_ and Δ*t*_*n*_ are common metrics in clinical studies of PSA dynamics after EBRT. According to our modeling framework, a high PSA nadir or a short time to reach it after EBRT conclusion are predictive for biochemical relapse (see Figure 3 and Table S1). Indeed, previous clinical studies have also linked these observations to worse prognosis, such as a higher likelihood of tumor recurrence and reduced patient survival [Zumsteg et al., 2015; Ray et al., 2006; Freiberger et al., 2017; Wein et al., 2012]. The estimation of *P*_*n*_ and Δ*t*_*n*_ with our models relies on *ρ*_*s*_ and *β*, but also on other model quantities that are not significantly different between non-relapsing and relapsing patients (see STAR Methods). This may partially explain their comparatively poorer performance in identifying biochemical relapse with respect to *ρ*_*s*_ and *β* [Lorenzo et al., 2019b]. Additionally, the post-EBRT PSA doubling time during biochemical relapse, a PSA metric linked to a poor PCa prognosis [Freiberger et al., 2017; Zumsteg et al., 2015; Bates et al., 2005; Wein et al., 2012], can also be estimated by leveraging our models [Lorenzo et al., 2019b].

Therefore, we believe that our model-based biomarkers could be leveraged to assess the probability of tumor recurrence and estimate patient survival after EBRT, thereby assisting the treating physician in therapeutic decision-making. We plan to explore these important applications of our modeling technology in future studies over larger cohorts. Additionally, in this study, we further investigate the performance of our model-based biomarkers to early identify biochemical relapse in the course of post-EBRT PSA monitoring. Our results show that the estimation of *ρ*_*s*_, *β, P*_*n*_, and Δ*t*_*n*_ with a fraction of the total PSA values available for each patient also enables to identify relapsing patients (see Figure 6 and Table 3). In general, we observe a similar classifier performance as in the global fitting scenario (see Figure 4 and Table 2), although a more conservative cutoff value may be required to optimally detect biochemical relapse with *ρ*_*s*_ (see Figure 6 and Table 3). The promising early classifier performance reported in this study suggests that our model-based biomarkers are robust with respect to the amount of data required to identify biochemical relapse, and thus may enable to anticipate the diagnosis of this event with respect to current methods relying on PSA threshold criteria (e.g., nadir+2 ng/mL). In this work, we perform a preliminary investigation of this hypothesis revealing that *ρ*_*s*_, *β*, and *P*_*n*_ may enable to detect biochemical relapse a median of 125 to 971.5 days (or equivalently 4.2 to 32.4 months), significantly outperforming the standard clinical practice. Interestingly, while *P*_*n*_ exhibits a poorer classifier performance than *ρ*_*s*_ and *β* according to the ROC curve analyses in this study (see Table 2 and Table 3), this biomarker rendered the earliest detection of biochemical relapse in our cohort. Future studies over larger cohorts of relapsing patients are necessary to precisely assess the predictive power of our model-based biomarkers and their combined ability to anticipate the detection of biochemical relapse with respect to standard practice.

### A flexible modeling framework to address practical and research applications

Regarding the comparative performance of the periodic and of the single dose model, this study does not provide conclusive results to select one over the other. First, both models show a remarkable agreement during fitting and forecasting PSA (e.g., see Figures 2 and 5). Despite the significantly superior fitting and forecasting results obtained with the single dose model for each patient in the non-relapsing subgroup, this difference is marginal in absolute terms (e.g., see Figures 2 and 5, Figure S1, and Tables S2 and S3). This observation was also confirmed when a comparison of the overall distributions of fit quality and prediction metrics did not identify significant differences in neither patient subgroup in the global fitting and the fitting-forecasting studies. Additionally, no significant differences were found in the fitting and forecasting of PSA values in the relapsing subgroup. Second, both models provide the same biomarkers of biochemical relapse, which exhibit comparatively equivalent classifier performance and predictive ability. This is demonstrated in the ROC analysis of the global fitting and the fitting-forecasting results as well as in the analysis of the DGBRD metric (see Figures 4, 6, and 7 along with Tables 2, 3, and 4). In our previous study [Lorenzo et al., 2019b], we highlighted that the single dose model exhibited an extraordinary performance despite its simplicity, which we observe again in the present work. Based on the similar performance of both models, we think that the simplicity of the single dose model makes it more amenable to facilitate the fitting and forecasting of PSA dynamics in real clinical scenarios. However, since the periodic dose model and the general parent formulation from which our models are derived account for the timing of radiation doses (see STAR Methods) [Lorenzo et al., 2019b], these modeling options may also be of interest to investigate the personalization of radiation plans (e.g., dose escalation, hypofractionation) [Mottet et al., 2021]. We provide additional comments on this point in the forthcoming section on the limitations of this study.

### Towards a robust clinical implementation

Around 20% to 50% of PCa patients undergoing radiotherapy are estimated to exhibit a biochemical relapse within 5 to 10 years of treatment conclusion [Kupelian et al., 2006; Rosenbaum et al., 2004]. Their early identification and the accurate estimation of the severity of their tumor recurrence is crucial to optimize disease control and survival. Further developments of our patient-specific forecasting methods based on mechanistic models of PSA dynamics could constitute a robust enabling technology to accurately address those timely needs in post-treatment patient monitoring. In particular, casting our models in a Bayesian framework would advance the current state of our forecasting technology by incorporating the uncertainty in PSA values [Carobene et al., 2018; Christensson et al., 2011] as well as the uncertainty emanating from the model parameterization and ensuing predictions [Lima et al., 2017; Hawkins-Daarud et al., 2019; Lorenzo et al., 2021]. We plan to investigate this approach to seamlessly integrate our mechanistic model predictions with uncertainty quantification and risk assessment. This strategy has the potential to guide clinical decision-making during the post-treatment monitoring of individual patients, including the frequency of PSA data collection, the timing of tests to ascertain the suspicion of tumor recurrence, the estimation of clinical risks and survival (e.g., biochemical relapse, local and distant recurrence, or death), and the planning of optimal primary and salvage treatments by maximizing therapeutic outcomes and minimizing toxicities.

### Limitations of the study

While the work presented herein shows promises for the use of mechanistic models of PSA dynamics to assist decision-making in post-EBRT monitoring of PCa, it also features some limitations that we plan to address in forthcoming studies. First, the patient cohort used for our analysis has a reduced number of relapsing patients (*n* = 10). Thus, to obtain a more robust analysis of the predictive performance of our mechanistic models and biomarkers, we need to extend our cohort to increase the number of patients showing biochemical relapse. This cohort extension would also provide enough statistical power to examine the correlations between our proposed model-based biomarkers and usual PCa clinical characteristics (e.g., Gleason score, TNM stage). To facilitate this effort, we plan to pool cohorts from multiple centers, which would also enable us to assess the applicability of our models across various institutions. Indeed, we believe that this is a key step towards the future clinical use of our predictive technology. Additionally, our analysis was focused on biochemical relapse detection as a surrogate for PCa recurrence [Mottet et al., 2021; Cornford et al., 2021; Wein et al., 2012]. To address this limitation, we specifically aim at extending our cohort with patients for whom PCa recurrence has been confirmed, including the type of recurrence (e.g., local or metastatic), which would let us analyze their correlation with our biomarkers and model predictions.

Second, all patients in the cohort leveraged in this study only had a single pre-EBRT PSA measurement. This limitation probably hindered the precise identification of baseline tumor cell dynamics by rendering tumor cell proliferation rates (*ρ*_*s*_) close to the minimal admissible value (see STAR Methods) in the non-relapsing subgroup and in the fitting-forecasting scenarios where observed PSA dynamics do not provide sufficient information to detect a rising branch (e.g., see Table S1 and [Lorenzo et al., 2019b]). However, this issue is compatible with our models to represent the early PSA decay usually observed in most patients after EBRT conclusion as well as the long-term plateauing PSA dynamics in cured patients [Zagars and Pollack, 1997; Cox et al., 1994; Hanlon et al., 1998; Vollmer and Montana, 1999; Taylor et al., 2005; Lorenzo et al., 2019b]. Thus, we think that underestimation of the tumor cell proliferation rate did not impede the performance of our models and biomarkers to respectively predict PSA dynamics and biochemical relapse, as shown by the results presented herein. Further pre-EBRT PSA values would specifically facilitate the estimation of the PCa cell proliferation rate, which may also lead to a more reliable estimation of the other model parameters. This improvement would refine our model and biomarker predictions, while also enabling the calculation of a surrogate for the proliferation activity of the tumor enabling the assessment of prognostic risks and expected survival [Cowen et al., 2002; Li et al., 2004; Berlin et al., 2017]. Moreover, the availability of several PSA values before EBRT may even allow to consider a different proliferation rate before and after EBRT (see STAR Methods). This modeling feature would enable the investigation of EBRT-mediated changes in tumor cell population dynamics, for example, due to direct changes to tumor cell cycle distribution and proliferation rates [Lima et al., 2017; Hormuth et al., 2018; Powathil et al., 2013] or due to the evolutionary treatment-induced promotion of a radiation-resistant, proliferative tumor phenotype [Greaves and Maley, 2012; Enriquez-Navas et al., 2015; Forouzannia et al., 2018; West et al., 2018].

Finally, our models assume that all PSA changes after EBRT conclusion emanate from radiation-induced tumor cell death and the proliferation of a potential tumor cell survival fraction. However, the mathematical formulation of our models could be extended to accommodate other mechanisms underlying PSA dynamics after EBRT. For instance, post-radiotherapy PSA bounces [Wein et al., 2012; Pinkawa et al., 2010; Freiberger et al., 2017] have been described *via* mechanistic modeling of the interplay between tumor cell dynamics and tumor immune response [Yamamoto et al., 2016]. Indeed, a recent study has also found this complex interplay to be central for the prediction of the probability of radiocurability of cancer patients [Alfonso et al., 2021]. Our models could also accommodate the increase in PSA caused by prostatic enlargement due to concomitant benign prostatic hyperplasia by means of an additional disease-specific PSA production term [Lorenzo et al., 2019b,a; Hanlon et al., 1998; Swanson et al., 2001], which could be informed by either population-based, age-stratified estimates of PSA changes due to this pathology [Roehrborn et al., 2000] or longitudinal patient-specific, imaging measurements of prostatic whole or central gland volumes [Lorenzo et al., 2019b,a; Cao et al., 2017; Roehrborn et al., 2000; Lieber et al., 2010]. Additionally, our mechanistic models of post-EBRT dynamics feature a model-naïve formulation of the survival fraction as a free parameter that is directly estimated from PSA data (see STAR Methods). This definition of the survival fraction could be refined by leveraging a radiobiological dose-dependent formulation, for which there exists a rich literature [Forouzannia et al., 2018; Corwin et al., 2013; Lima et al., 2017; Bodgi et al., 2016; Rockne et al., 2015; Powathil et al., 2007, 2013; Lewin et al., 2018; O’Rourke et al., 2008; Kal and Gellekom, 2003; Wang and Li, 2005]. Hence, our modeling framework would enable to investigate alternative radiation plans and systematically select clinically-feasible, optimal regimens for individual patients [Forouzannia et al., 2018; Henares-Molina et al., 2017; Brüningk et al., 2021; Lipková et al., 2019; Ayala-Hernández et al., 2021]. The aforementioned model extensions increase the number of parameters to be identified on a patient-specific basis. Thus, global sensitivity analysis and model selection would be recommendable to identify a minimal set of driving parameters requiring personalized calibration and assess whether the extended models are superior to the parent formulations used in this study [Oden et al., 2016; Lorenzo et al., 2021].

## Data Availability

De-identified data and all original code are available online at Zenodo (https://doi.org/10.5281/zenodo.6277674).

## STAR Methods

### Key resources table

**Table 5.** Key resources table

| Resource | Source | Identifier |
| --- | --- | --- |
| **Deposited data** |  |  |
| De-identified patient data | This paper | https://doi.org/10.5281/zenodo.6277674 |
| **Software and algorithms** |  |  |
| MATLAB R2021a | The Mathworks (Natick, MA, USA) | https://www.mathworks.com/products/matlab.html |
| MATLAB scripts for model fitting and forecasting | This paper | https://doi.org/10.5281/zenodo.6277674 |
| MATLAB scripts for statistical analysis | This paper | https://doi.org/10.5281/zenodo.6277674 |

### Resource availability

#### Lead contact

Further information and requests for resources should be directed to the lead contact, Guillermo Lorenzo.

#### Materials availability

This study did not generate new unique reagents.

#### Data and code availability

De-identified patient data and all original code have been deposited at Zenodo (https://doi.org/10.5281/zenodo.6277674) and are publicly available as of the date of publication. Any additional information required to reanalyze the data reported in this paper is available from the lead contact upon request.

### Subject details

Anonymized patient data were retrospectively collected at Istituto di Ricovero e Cura a Carattere Scientifico Ospedale San Raffaele (IRCCSOSR, Milan, Italy). Ethics approval and informed consent waiver were obtained from the Internal Review Board at IRCCSOSR for this study. The inclusion criteria were diagnosis of localized or locally advanced PCa (clinical TNM stage: T1 to T3, not N1 or M1), availability of complete basic diagnostic data (i.e., baseline PSA, Gleason score, and TNM stage), EBRT as primary treatment with curative intent and without (neo)adjuvant ADT, follow-up for at least 3 years since the onset of radiotherapy, and a PSA history featuring at least 5 values after EBRT conclusion. The exclusion criteria were a previous diagnosis of cancer prior to PCa, any other prior or concomitant treatment for PCa (e.g., ADT, radical prostatectomy, radiotherapy, chemotherapy), EBRT without curative intent, incomplete diagnostic data, and insufficient PSA monitoring for this study.

A total of 206 men treated with EBRT at IRCCS Ospedale San Raffaele between years 2006 and 2018 were initially considered for this study. The application of the inclusion and exclusion criteria resulted in a final cohort of 166 patients. A total of 10 patients in this cohort were diagnosed with biochemical relapse and tumor recurrence was confirmed in five of them with choline PET/CT. PCa recurrence was local in one patient and metastatic in the other four patients. Nine relapsing patients received a secondary treatment, which consisted of ADT for seven patients, radiotherapy in one patient, and combined ADT and radiotherapy for another patient. The PSA data collected after the onset of the second treatment is not considered in the present study. We recall that Table 1 summarizes the characteristics of the study cohort, the relapsing subgroup, and the non-relapsing subgroup. The total number of PSA values reported for each patient (*n*_*P*_) includes a single pre-EBRT value (i.e., the baseline PSA, *P*_*d*_) and the series of PSA values collected during post-EBRT follow-up. A Wilcoxon rank-sum test identified significantly larger PSA at diagnosis (*P*_*d*_, *p* = 0.017), higher number of PSA values (*n*_*p*_, *p* = 0.040), and more frequent PSA testing (*p <* 0.001) in the relapsing subgroup. Additionally, the proportion of T1, T2, and T3 disease in the non-relapsing/relapsing subgroups are 88/6, 64/4, and 4/0, respectively.

### Method details

#### General mathematical model

We call *P*(*t*) the serum PSA at time *t*. Our time interval of interest is (*t*_0_,*t* _*f*_), where *t*_0_ is the time at which the pre-EBTR PSA measurement in our database was taken, and *t* _*f*_ is the latest time at which we forecast the PSA. For simplicity, we rescale time such that *t*_0_ = 0 for all patients. The patients in our database received *n*_*d*_ radiation doses at times 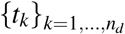, where 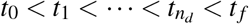. Note that the *t*_*k*_’s may vary from patient to patient.

We assume that the serum PSA is proportional to the number of tumor cells *N*(*t*), that is

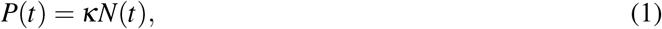

where *κ* is a constant. Prior to EBRT treatment, we assume that *N* grows exponentially in time from *N*(*t*_0_) = *N*_0_, at a characteristic rate *ρ*_*n*_. Thus,

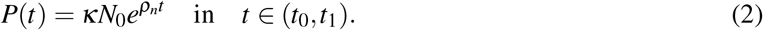

From Eq. (2), we define *P*_0_ = *P*(0) = *κN*_0_.

For each time interval, *I*_*k*_ = (*t*_*k*_,*t*_*k*+1_), *k* = 1, …, *n*_*d*_ − 1, we define *S*_*k*_(*t*), which represents the fraction of tumor cells surviving to the *k*-th radiation dose, and 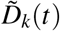, which is the fraction of tumor cells irreversibly damaged after the *k*-th radiation dose. For compactness of the notation, we also define *S*_0_(*t*) and 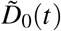 in the interval (*t*_0_,*t*_1_) as *S*_0_(*t*) = *N*(*t*) and 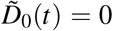. This assumes that there are no damaged cells before treatment.

The values of *S*_*k*_ and 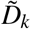 at time *t*_*k*_ are obtained from 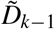 and *S*_*k*−1_ as

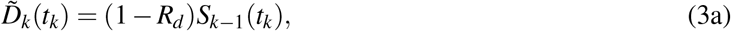

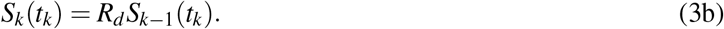

Eq. (3a) assumes that the radiation dose at time *t*_*k*_ immediately produces irreversible damage to a fraction of cells (1 − *R*_*d*_), where 0 *< R*_*d*_ *<* 1. The remaining fraction of cells, *R*_*d*_, continues in the compartment of surviving cells. The parameter *R*_*d*_ is patient specific, but constant for all doses. We do not assume any specific formulation for *R*_*d*_, but simply compute it from the PSA data. Eqs. (3a)–(3b) provide initial conditions for the ordinary differential equations (ODEs) that govern the dynamics of 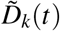 and *S*_*k*_(*t*) in the time interval *I*_*k*_. These ODEs are based on the assumptions that irreversibly damaged cells undergo programmed cell death at a rate *ρ*_*d*_, and surviving cells continue their proliferation at a characteristic rate *ρ*_*s*_,

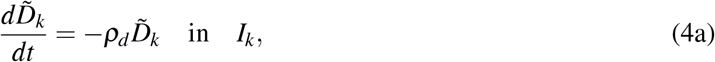

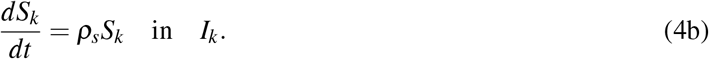

We further define the cumulative number of irreversibly damaged tumor cells in the time interval *I*_*k*_ as

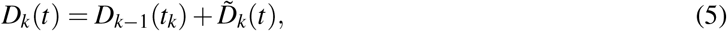

with *D*_0_(*t*) = 0. Hence, the total population of tumor cells in the interval *I*_*k*_ is

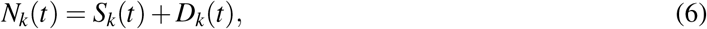

and the PSA is given by

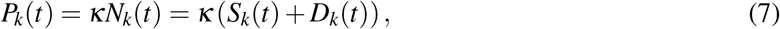

Note that Eqs. (3a)–(4b) can be solved recursively on all time intervals from *I*_1_ to 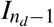 through direct integration, which leads to

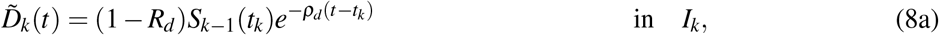

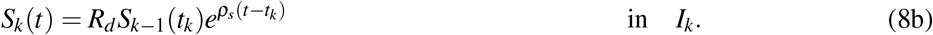

From Eqs. (8a) and (5), we obtain

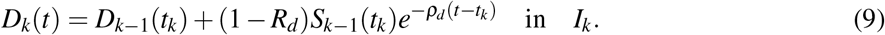

By applying Eqs. (8b) and (5) recursively, we derive

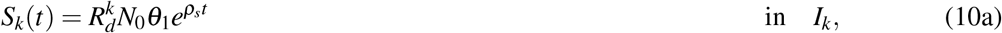

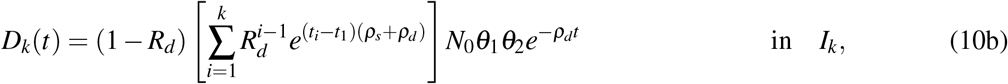

where 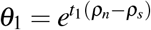 and 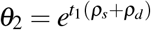. Then, the expression for the serum PSA is

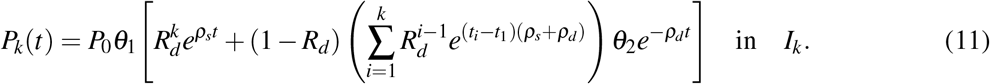

#### Periodic dose model

In the periodic dose model, we assume that the radiation doses are evenly spaced in time with a constant intertreatment interval *τ*_*r*_, i.e., *t*_*k*_ = *t*_1_ + (*k* − 1)*τ*_*r*_ for *k* = 1, …, *n*_*d*_. This allows us to simplify Eq. (10b) into

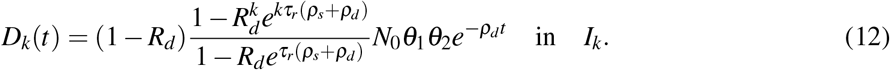

Then, the PSA is given by

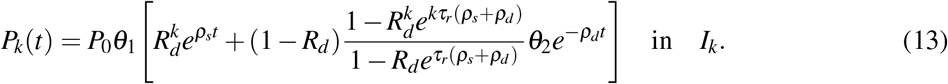

#### Single dose model

In the single dose model, we assume that the entire radiation treatment is given in one single dose at time *t*_*D*_. Then,

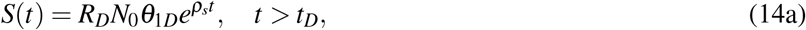

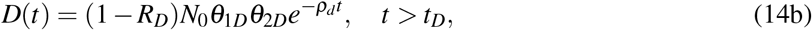

where *R*_*D*_ is the fraction of surviving tumor cells after the entire treatment dose, 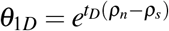 and 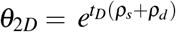. Then, the PSA is given by

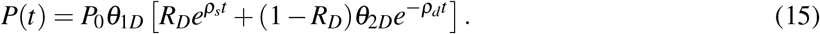

#### Dimensional analysis and predicted PSA nadir

The PSA dynamics after treatment completion can be obtained from Eq. (13) as

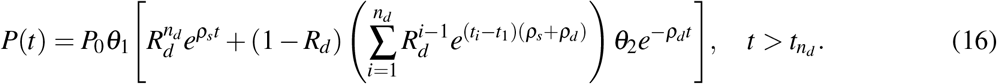

We nondimensionalize PSA and time using the scales 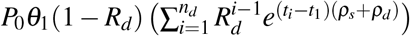 and 1*/ρ*_*d*_, respectively. Then, using hats to denote dimensionless quantities, we have

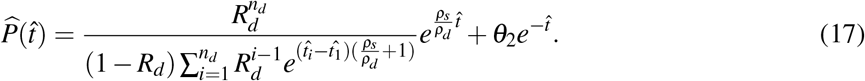

Defining the nondimensional groups

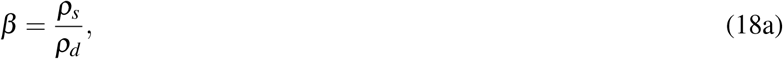

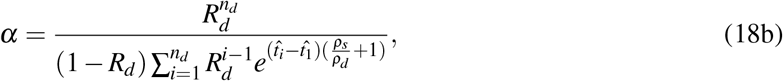

we can express the dimensionless PSA as

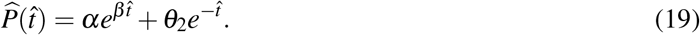

The parameter *α* represents the efficacy of the radiation plan and *β* defines the dynamics of the tumor cell population after radiation. While *β* is independent from the treatment plan, *α* can be specialized to the two treatment plans studied in this paper. For the periodic dose treatment

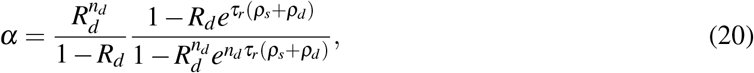

while for the single dose treatment

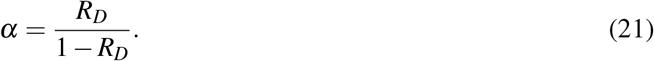

The nondimenzionalized PSA velocity is defined as

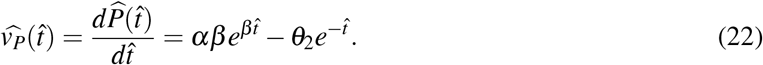

The PSA nadir is achieved at a dimensionless time 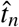 defined by 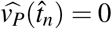. Using Eq. (22), we find

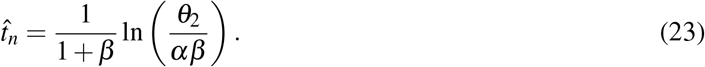

The corresponding dimensional time is

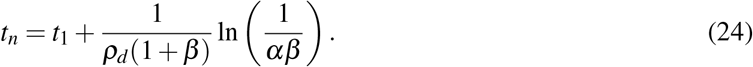

and the time to PSA nadir since the completion of the treatment is given by 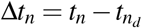.

#### Further modeling assumptions

Some patient may experience delays in their radiation plans due to treatment side effects, local holidays, or machine routine maintenance. Additionally, the patient dataset used in this study only includes the times of EBRT initiation and conclusion, the radiation dose, and the number of doses. This information is not compatible with an accurate use of our general model, which would require the exact dates of each EBRT fraction for each patient. To overcome this limitation, we introduced the assumptions on radiation delivery that lead to the periodic and single dose models presented above. Thus, here we perform our analyses with these two models, which can be leveraged as surrogates of the general model as shown in [Lorenzo et al., 2019b].

We further assume that EBRT does not change the proliferation rate of the surviving tumor cells, such that *ρ*_*n*_ = *ρ*_*s*_ and *θ*_1_ = *θ*_1*D*_ = 1. This is a common assumption in the literature [Lima et al., 2017; Corwin et al., 2013; Pérez-García et al., 2015] that facilitates the parameterization of our models using the cohort of this study, which only features one pre-EBRT PSA value to inform *ρ*_*n*_. Additionally, we set *t*_*D*_ = *t*_1_ for the single dose model as in [Lorenzo et al., 2019b].

#### Model fitting and forecasting

We fit the PSA data from each patient to both the periodic and single dose models. We perform model fitting by leveraging a nonlinear least-squares method based on a trust-region reflective algorithm. Table 6 provides the initial guess as well as the lower and upper bounds of the model parameters for both the periodic and the single dose models. Our model fitting method aims at minimizing an objective functional *J*, given by

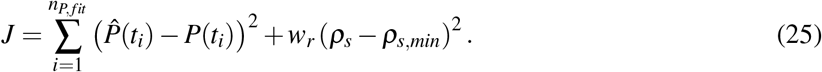

**Table 6.** Parameter initial values and bounds for model fitting. *P*_*d*_ is the baseline PSA value for each patient (see Table 1).

| Parameter | Units | Initial value | Lower bound | Upper bound |
| --- | --- | --- | --- | --- |
| $P_0$ | (ng/mL) | $P_d$ | 0 | 100 |
| $R_d$ | - | 0.9 | 0.5 | 1 |
| $R_D$ | - | $0.9^{n_d}$ | $0.5^{n_d}$ | 1 |
| $\rho_s$ | (1/mo) | 0.02 | 0.001 | 2 |
| $\rho_d$ | (1/mo) | 0.5 | 0.001 | 2 |

The first term in the right-hand side of Eq. (25) represents the mismatch between PSA data 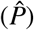 and the model estimation of PSA (*P*) at each of the PSA test times *t*_*i*_ (*i* = 1, …, *n*_*P,fit*_). For each model and patient, we run a total of *n*_*P,fit*_ model fits in the fitting-forecasting study (*n*_*P,fit*_ = 5, …, *n*_*P*_ − 1), whereas we only perform a single model fit with *n*_*P,fit*_ = *n*_*P*_ in the global fitting study. The second term in the right-hand side of Eq. (25) regularizes the proliferation rate of the surviving tumor cells (*ρ*_*s*_) to low values and was introduced to limit overfitting, especially when only a small number of PSA values were available to calculate the model parameters. The regularization weight *w*_*r*_ is empirically set at 2500 and *ρ*_*s,min*_ is the minimum admissible value for this parameter in Table 6.

We further cast our model fitting problem within a multi-start strategy to facilitate convergence for all patient datasets and avoid the selection of local minima in the minimization problem outlined above. In brief, this strategy consists of solving the model fitting problem multiple times, each of them using a different initial guess and resulting in a parameter set if convergence is achieved. Then, the algorithm selects the resulting parameter set that renders the lowest value of the objective functional as the global minimum. We use a collection of 20 initial guesses that are randomly sampled within the parameter space and always includes the one provided in Table 6.

#### Code implementation

The calculations based on the numerical methods described in this section are performed using MATLAB (R2021a, The Mathworks, Natick, MA, USA). In particular, model fitting is implemented by leveraging the Global Optimization Toolbox.

### Quantification and statistical analysis

#### Quantitative assessment of model fits and forecasts

In the global fitting study, the quality of fit is assessed by means of the root mean squared error (RMSE) and the coefficient of determination (*R*^2^). Given that some *n*_*P,fit*_ scenarios involved a reduced number of PSA values to assess the model forecasts, we only use the RMSE to analyze the quality of fit and validate the model predictions of PSA dynamics in the fitting-forecasting study. Additionally, we calculate the 95% nonlinear regression prediction confidence intervals for our model fits and forecasts.

#### Receiver operating characteristic curves

We calculate the receiver operating characteristic (ROC) curves for our model-based biomarkers obtained from global fitting and the fitting-forecasting study with either PSA model. Global fitting produces a unique value for each model-based biomarker per patient. We use the resulting set of values obtained across the whole cohort as the thresholds to construct the corresponding ROC curves. Since all PSA values for each patient are used in global fitting, these ROC curves assess the ability of the model-based biomarkers to retrospectively classify patients as relapsing or non-relapsing. The fitting-forecasting study produces a set of values for each model-based biomarker per patient, which result from the sequential model fits to an increasing number of PSA values ranging from *n*_*P,fit*_ = 5 to *n*_*P,fit*_ = *n*_*P*_ − 1. Hence, each set contains the temporal evolution of each model-based biomarker during post-EBRT monitoring for each patient. To construct the ROC curve of each model-based biomarker in the fitting-forecasting study, we first pool all the biomarker values across all patients to define the thresholds. Then, for each patient, we assess whether each threshold can identify any of the biomarker values obtained across the *n*_*P,fit*_ scenarios as predictive for biochemical relapse. Thus, in this scenario, the ROC curves provide an assessment of the ability of the model-based biomarkers to early classify the patients as relapsing or non-relapsing during the course of post-EBRT PSA monitoring.

For each ROC curve, we further calculate the area under the curve (AUC) using the trapezoidal rule and the optimal performance point according to Youdens index. Additionally, we calculate the 95% bootstrap confidence intervals of the ROC curves and their corresponding AUC and optimal performance point. We use 2000 bootstrap samples for these calculations. The 95% bootstrap confidence interval for each ROC curve is obtained as the envelope of the 95% bootstrap confidence interval regions obtained for each threshold value used in the construction of the ROC curve along the sensitivity and specificity axes.

#### Statistical analysis

We use Wilcoxon rank-sum and signed-rank tests for the statistical analyses performed in this work. In the Results section, we specify when we use each type of test and whether it is two-tailed or one tailed for each statistical analysis of the global fitting and fitting-forecasting study results. The level of significance for all statistical tests is set at 5%.

#### Code implementation

The calculations based on the statistical methods described in this section are performed using MATLAB (R2021a, The Mathworks, Natick, MA, USA). In particular, we use the Statistics and Machine Learning Toolbox to calculate the 95% nonlinear regression prediction confidence intervals for our model fits and forecasts, construct the 95% bootstrap confidence intervals for the ROC curve analysis, and perform the aforementioned statistical tests.

## Acknowledgments

This project has received funding from the European Union’s Horizon 2020 research and innovation programme under the Marie Skłodowska-Curie grant agreement No. 838786. A.R. was partially supported by the MIUR-PRIN project XFAST-SIMS (No. 20173C478N). H.G. was partially funded by the Purdue Center for Cancer Research through a Concept Grant. V.M.P.G. is partially supported by the Spanish Ministerio de Ciencia e Innovación (grant PID2019-110895RB-100, doi:10.13039/501100011033).

## Author contributions

Conceptualization: G.L., V.M.P.G., H.G., A.R.

Methodology: G.L., V.M.P.G., H.G., A.R.

Software: G.L.

Validation: G.L.

Formal Analysis: G.L., V.M.P.G., H.G., A.R.

Investigation: G.L., N.M., V.M.P.G., H.G., A.R.

Resources: N.M., C.L.D., C.C., A.F., A.B, F.M.

Data Curation: G.L., N.M., C.L.D., C.C., A.F., A.B., F.M.

Writing - Original Draft Preparation: G.L., N.M., V.M.P.G., H.G., A.R.

Writing - Review & Editing: G.L., N.M., C.L.D., C.C., A.F., A.B., F.M., V.M.P.G., H.G., A.R.

Visualization: G.L.

Supervision: G.L., A.R.

Project Administration: G.L.

Funding Acquisition: G.L., A.R.

## Supplemental Information

**Table S1.** Distribution of model-based quantities of interest obtained in the global fitting study using the periodic dose and the single dose models. This table complements Figure 3 in the main text. IQR: interquartile range.

| QOI | All patients ($n=166$) | | | Non-relapsing patients ($n=156$) | | | Relapsing patients ($n=10$) | | |
| --- | --- | --- | --- | --- | --- | --- | --- | --- | --- |
|  | Median | IQR | Range | Median | IQR | Range | Median | IQR | Range |
| **Periodic dose model** |  |  |  |  |  |  |  |  |  |
| $P_0$ (ng/mL) | 6.4 | (4.6, 8.8) | (0.6, 81.0) | 6.2 | (4.5, 8.3) | (0.6, 20.5) | 9.6 | (5.9, 13.1) | (5.0, 81.0) |
| $R_d$ | 0.89 | (0.88, 0.91) | (0.80, 0.98) | 0.89 | (0.88, 0.91) | (0.80, 0.98) | 0.88 | (0.84, 0.91) | (0.80, 0.94) |
| $\rho_d$ (1/mo) | 0.31 | (0.22, 0.47) | (0.05, 2.0) | 0.30 | (0.22, 0.47) | (0.05, 2.0) | 0.35 | (0.17, 0.39) | (0.06, 2.0) |
| $\rho_s$<br>($10^{-3}$/mo) | 1.0 | (1.0, 1.0) | (1.0, 46.8) | 1.0 | (1.0, 1.0) | (1.0, 13.9) | 17.6 | (16.0, 20.1) | (14.6, 46.8) |
| $\beta$ ($10^{-3}$) | 3.6 | (2.3, 5.2) | (0.5, 802.5) | 3.5 | (2.2, 4.8) | (0.5, 75.6) | 51.2 | (37.1, 133.8) | (9.0, 802.5) |
| $\alpha$ ($10^{-2}$) | 3.6 | (2.3, 6.8) | (0.1, 29.6) | 3.7 | (2.4, 6.8) | (0.1, 29.6) | 2.3 | (0.6, 6.6) | (0.2, 15.6) |
| $P_n$ (ng/mL) | 0.3 | (0.2, 0.5) | (0.0, 1.8) | 0.3 | (0.2, 0.5) | (0.0, 1.8) | 0.8 | (0.6, 1.0) | (0.3, 1.3) |
| $\Delta t_n$ (mo) | 26.8 | (17.5, 36.6) | (3.1, 142.7) | 27.1 | (19.3, 37.4) | (3.1, 142.7) | 16.5 | (10.9, 33.8) | (3.6, 50.8) |
| **Single dose model** |  |  |  |  |  |  |  |  |  |
| $P_0$ (ng/mL) | 6.4 | (4.6, 8.8) | (0.6, 81.0) | 6.2 | (4.5, 8.3) | (0.6, 20.7) | 9.6 | (5.9, 13.1) | (5.0, 81.0) |
| $R_D$ ($10^{-2}$) | 4.0 | (2.6, 6.9) | (0.0, 55.1) | 4.0 | (2.6, 7.0) | (0.0, 55.1) | 2.5 | (0.8, 6.7) | (0.2, 16.0) |
| $\rho_d$ (1/mo) | 0.29 | (0.21, 0.43) | (0.05, 2.0) | 0.29 | (0.21, 0.43) | (0.05, 2.0) | 0.32 | (0.17, 0.37) | (0.06, 2.0) |
| $\rho_s$<br>($10^{-3}$/mo) | 1.0 | (1.0, 1.0) | (1.0, 47.5) | 1.0 | (1.0, 1.0) | (1.0, 13.9) | 17.7 | (16.3, 21.0) | (14.7, 47.5) |
| $\beta$ ($10^{-3}$) | 3.9 | (2.5, 5.5) | (0.5, 838.0) | 3.7 | (2.4, 5.1) | (0.5, 76.1) | 60.6 | (39.3, 138.9) | (9.0, 838.0) |
| $\alpha$ ($10^{-2}$) | 4.2 | (2.6, 7.4) | (0.0, 122.5) | 4.2 | (2.7, 7.5) | (0.0, 122.5) | 2.6 | (0.8, 7.2) | (0.2, 19.1) |
| $P_n$ (ng/mL) | 0.3 | (0.2, 0.5) | (0.0, 1.7) | 0.3 | (0.2, 0.4) | (0.0, 1.7) | 0.8 | (0.6, 1.0) | (0.3, 1.3) |
| $\Delta t_n$ (mo) | 28.6 | (18.9, 38.8) | (2.4, 384.8) | 29.2 | (20.6, 39.2) | (2.4, 384.8) | 17.0 | (11.8, 34.6) | (3.2, 51.4) |

**Figure S1.**
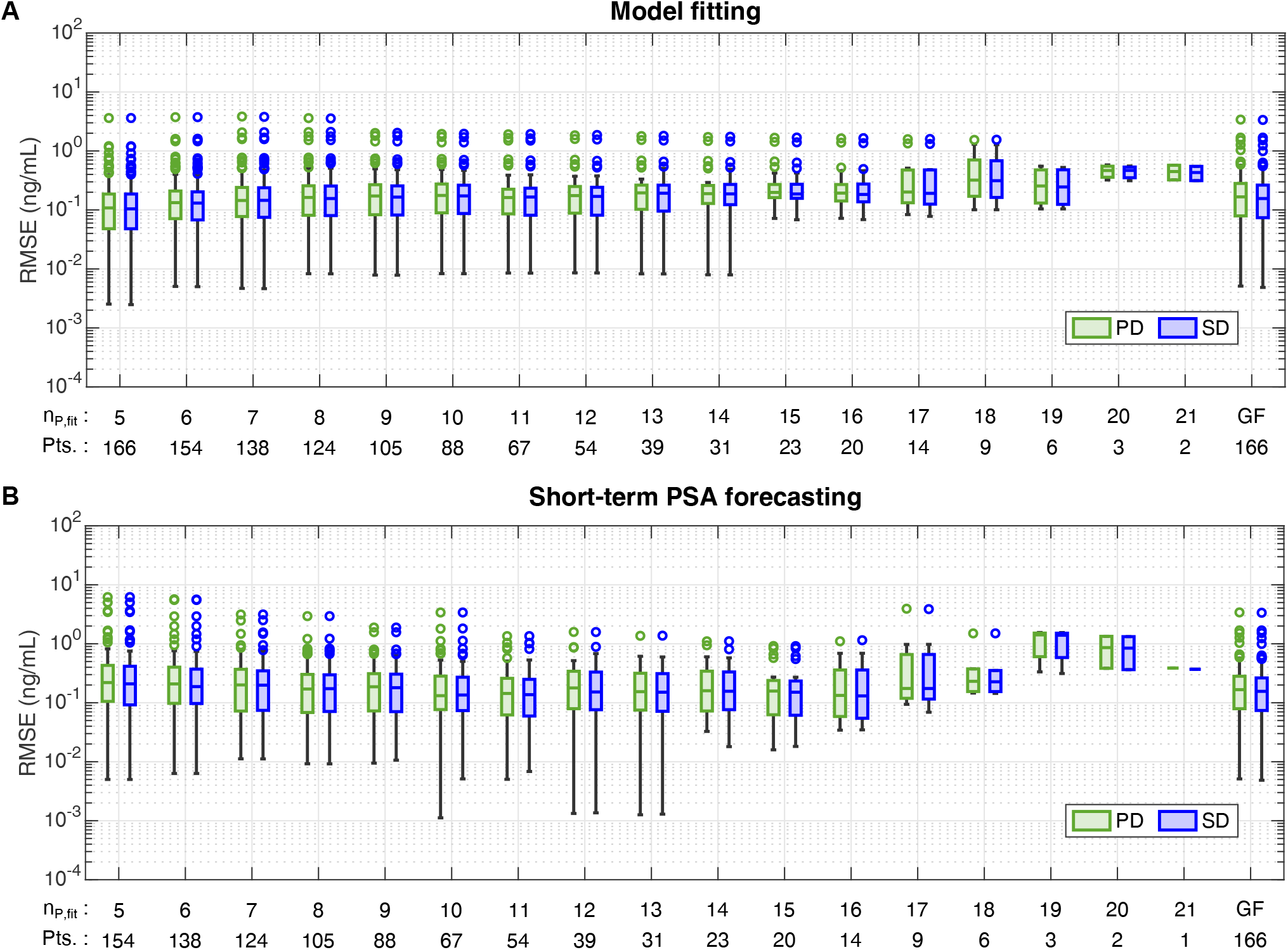
Distributions of the RMSE of model fitting and short-term PSA forecasting across the *n*_*P,fit*_ scenarios of the fitting-forecasting study. Panel (A) shows the boxplots of the RMSE (ng/mL) obtained in the fitting of the mechanistic models of PSA dynamics to *n*_*P,fit*_ = 5, …, 21 PSA values. Panel (B) shows the boxplots of the RMSE (ng/mL) obtained in the short-term prediction of 2 PSA values using the mechanistic models of PSA dynamics fitted to *n*_*P,fit*_ = 5, …, 21 PSA values. For each individual patient *n*_*P,fit*_ = 5, …, *n*_*P*_ − 1, where *n*_*P*_ is the number of PSA values available for each of them (see Table 1). The horizontal axis in each panel features the number of patients (Pts.) included in each *n*_*P,fit*_ scenario. The last boxplots in each panel further provide the RMSE distribution obtained during global fitting for each model (GF) as a reference of the ultimate RMSE value when the models are informed with all available PSA data from each patient. Green boxplots show results from the periodic dose model (PD), while blue boxplots correspond to the single dose model (SD). Outliers are represented as hollow circles. This figure is complemented by Tables S2 and S3.

**Table S2.** Distribution of the RMSE values (ng/mL) resulting from fitting the periodic and single dose models to *n*_*P,fit*_ PSA values in the fitting-forecasting study. This table complements Figure S1. IQR: interquartile range.

| $n_{p,fit}$ | All patients | | | | Non-relapsing patients | | | | Relapsing patients | | | |
| --- | --- | --- | --- | --- | --- | --- | --- | --- | --- | --- | --- | --- |
| | $n$ | Median | IQR | Range | $n$ | Median | IQR | Range | $n$ | Median | IQR | Range |
| **Periodic dose model** |  |  |  |  |  |  |  |  |  |  |  |  |
| 5 | 166 | 0.11 | (0.05, 0.19) | (0.00, 3.62) | 156 | 0.11 | (0.05, 0.18) | (0.00, 3.62) | 10 | 0.16 | (0.09, 0.78) | (0.02, 1.21) |
| 6 | 154 | 0.13 | (0.07, 0.21) | (0.01, 3.74) | 144 | 0.13 | (0.07, 0.21) | (0.01, 3.74) | 10 | 0.18 | (0.10, 0.80) | (0.03, 1.60) |
| 7 | 138 | 0.14 | (0.08, 0.24) | (0.00, 3.84) | 129 | 0.14 | (0.08, 0.23) | (0.00, 3.84) | 9 | 0.17 | (0.10, 0.97) | (0.05, 2.07) |
| 8 | 124 | 0.16 | (0.08, 0.26) | (0.01, 3.61) | 115 | 0.16 | (0.08, 0.25) | (0.01, 3.61) | 9 | 0.19 | (0.12, 0.92) | (0.06, 2.09) |
| 9 | 105 | 0.17 | (0.08, 0.27) | (0.01, 2.02) | 97 | 0.17 | (0.08, 0.26) | (0.01, 1.80) | 8 | 0.18 | (0.14, 1.07) | (0.05, 2.02) |
| 10 | 88 | 0.18 | (0.09, 0.28) | (0.01, 1.95) | 81 | 0.18 | (0.09, 0.27) | (0.01, 1.73) | 7 | 0.22 | (0.13, 1.21) | (0.05, 1.95) |
| 11 | 67 | 0.16 | (0.09, 0.22) | (0.01, 1.92) | 60 | 0.16 | (0.08, 0.22) | (0.01, 1.66) | 7 | 0.27 | (0.17, 1.28) | (0.06, 1.92) |
| 12 | 54 | 0.18 | (0.09, 0.25) | (0.01, 1.84) | 48 | 0.16 | (0.08, 0.23) | (0.01, 1.60) | 6 | 0.28 | (0.18, 0.60) | (0.09, 1.84) |
| 13 | 39 | 0.19 | (0.10, 0.26) | (0.01, 1.79) | 34 | 0.19 | (0.09, 0.24) | (0.01, 1.53) | 5 | 0.27 | (0.21, 0.88) | (0.17, 1.79) |
| 14 | 31 | 0.19 | (0.13, 0.26) | (0.01, 1.73) | 27 | 0.18 | (0.10, 0.24) | (0.01, 1.48) | 4 | 0.42 | (0.25, 1.14) | (0.21, 1.73) |
| 15 | 23 | 0.20 | (0.16, 0.27) | (0.07, 1.67) | 19 | 0.18 | (0.14, 0.24) | (0.07, 1.43) | 4 | 0.48 | (0.31, 1.11) | (0.20, 1.67) |
| 16 | 20 | 0.19 | (0.14, 0.27) | (0.07, 1.63) | 16 | 0.18 | (0.13, 0.21) | (0.07, 1.38) | 4 | 0.49 | (0.34, 1.08) | (0.22, 1.63) |
| 17 | 14 | 0.20 | (0.13, 0.48) | (0.08, 1.59) | 11 | 0.19 | (0.12, 0.23) | (0.08, 1.34) | 3 | 0.51 | (0.48, 1.32) | (0.48, 1.59) |
| 18 | 9 | 0.32 | (0.17, 0.70) | (0.10, 1.54) | 6 | 0.19 | (0.13, 0.32) | (0.10, 1.30) | 3 | 0.51 | (0.49, 1.28) | (0.48, 1.54) |
| 19 | 6 | 0.26 | (0.13, 0.48) | (0.10, 0.55) | 4 | 0.16 | (0.12, 0.26) | (0.10, 0.33) | 2 | 0.51 | - | (0.48, 0.55) |
| 20 | 3 | 0.47 | (0.36, 0.55) | (0.32, 0.58) | 1 | 0.32 | - | - | 2 | 0.52 | - | (0.47, 0.58) |
| 21 | 2 | 0.45 | - | (0.32, 0.57) | 1 | 0.32 | - | - | 1 | 0.57 | - | - |
| Pooled | 1043 | 0.16 | (0.08, 0.25) | (0.00, 3.84) | 949 | 0.15 | (0.08, 0.23) | (0.00, 3.84) | 94 | 0.34 | (0.16, 0.76) | (0.02, 2.09) |
| **Single dose model** |  |  |  |  |  |  |  |  |  |  |  |  |
| 5 | 166 | 0.10 | (0.05, 0.19) | (0.00, 3.62) | 156 | 0.10 | (0.05, 0.18) | (0.00, 3.62) | 10 | 0.18 | (0.09, 0.74) | (0.01, 1.20) |
| 6 | 154 | 0.13 | (0.07, 0.20) | (0.01, 3.74) | 144 | 0.13 | (0.07, 0.20) | (0.01, 3.74) | 10 | 0.19 | (0.10, 0.75) | (0.02, 1.61) |
| 7 | 138 | 0.15 | (0.08, 0.24) | (0.00, 3.79) | 129 | 0.14 | (0.07, 0.21) | (0.00, 3.79) | 9 | 0.19 | (0.10, 0.93) | (0.04, 2.08) |
| 8 | 124 | 0.16 | (0.08, 0.25) | (0.01, 3.56) | 115 | 0.15 | (0.08, 0.25) | (0.01, 3.56) | 9 | 0.19 | (0.11, 0.89) | (0.04, 2.11) |
| 9 | 105 | 0.17 | (0.08, 0.26) | (0.01, 2.03) | 97 | 0.16 | (0.08, 0.25) | (0.01, 1.80) | 8 | 0.18 | (0.14, 1.05) | (0.04, 2.03) |
| 10 | 88 | 0.17 | (0.09, 0.26) | (0.01, 1.96) | 81 | 0.17 | (0.08, 0.25) | (0.01, 1.71) | 7 | 0.22 | (0.13, 1.20) | (0.04, 1.96) |
| 11 | 67 | 0.17 | (0.08, 0.23) | (0.01, 1.94) | 60 | 0.16 | (0.08, 0.22) | (0.01, 1.63) | 7 | 0.27 | (0.19, 1.27) | (0.05, 1.94) |
| 12 | 54 | 0.17 | (0.08, 0.24) | (0.01, 1.87) | 48 | 0.16 | (0.08, 0.23) | (0.01, 1.56) | 6 | 0.28 | (0.19, 0.56) | (0.09, 1.87) |
| 13 | 39 | 0.19 | (0.10, 0.26) | (0.01, 1.81) | 34 | 0.18 | (0.09, 0.23) | (0.01, 1.50) | 5 | 0.27 | (0.21, 0.86) | (0.17, 1.81) |
| 14 | 31 | 0.19 | (0.12, 0.27) | (0.01, 1.74) | 27 | 0.18 | (0.09, 0.24) | (0.01, 1.45) | 4 | 0.41 | (0.25, 1.13) | (0.21, 1.74) |
| 15 | 23 | 0.19 | (0.16, 0.27) | (0.07, 1.69) | 19 | 0.18 | (0.13, 0.23) | (0.07, 1.40) | 4 | 0.46 | (0.31, 1.10) | (0.21, 1.69) |
| 16 | 20 | 0.18 | (0.14, 0.28) | (0.07, 1.65) | 16 | 0.18 | (0.12, 0.21) | (0.07, 1.36) | 4 | 0.48 | (0.34, 1.07) | (0.22, 1.65) |
| 17 | 14 | 0.19 | (0.13, 0.48) | (0.08, 1.60) | 11 | 0.18 | (0.12, 0.24) | (0.08, 1.32) | 3 | 0.48 | (0.48, 1.32) | (0.48, 1.60) |
| 18 | 9 | 0.31 | (0.16, 0.68) | (0.10, 1.55) | 6 | 0.18 | (0.12, 0.31) | (0.10, 1.28) | 3 | 0.48 | (0.48, 1.28) | (0.48, 1.55) |
| 19 | 6 | 0.25 | (0.12, 0.48) | (0.10, 0.53) | 4 | 0.15 | (0.11, 0.25) | (0.10, 0.32) | 2 | 0.50 | - | (0.48, 0.53) |
| 20 | 3 | 0.47 | (0.35, 0.53) | (0.31, 0.56) | 1 | 0.31 | - | - | 2 | 0.51 | - | (0.47, 0.56) |
| 21 | 2 | 0.43 | - | (0.31, 0.55) | 1 | 0.31 | - | - | 1 | 0.55 | - | - |
| Pooled | 1043 | 0.15 | (0.08, 0.25) | (0.00, 3.79) | 949 | 0.14 | (0.07, 0.22) | (0.00, 3.79) | 94 | 0.34 | (0.17, 0.72) | (0.01, 2.11) |

**Table S3.** Distribution of the RMSE values (ng/mL) for a short-term forecast of two PSA values using the periodic and single dose models fitted to *n*_*P,fit*_ PSA values in the fitting-forecasting study. This table complements Figure S1. IQR: interquartile range.

| $n_{p,fit}$ | All patients | | | | Non-relapsing patients | | | | Relapsing patients | | | |
| --- | --- | --- | --- | --- | --- | --- | --- | --- | --- | --- | --- | --- |
| | $n$ | Median | IQR | Range | $n$ | Median | IQR | Range | $n$ | Median | IQR | Range |
| **Periodic dose model** |  |  |  |  |  |  |  |  |  |  |  |  |
| 5 | 154 | 0.22 | (0.11, 0.43) | (0.01, 6.19) | 144 | 0.21 | (0.09, 0.42) | (0.01, 6.19) | 10 | 0.52 | (0.25, 1.68) | (0.12, 3.62) |
| 6 | 138 | 0.21 | (0.10, 0.40) | (0.01, 5.69) | 129 | 0.21 | (0.09, 0.37) | (0.01, 5.53) | 9 | 0.58 | (0.14, 1.39) | (0.07, 5.69) |
| 7 | 124 | 0.20 | (0.07, 0.37) | (0.01, 3.14) | 115 | 0.20 | (0.07, 0.35) | (0.01, 3.14) | 9 | 0.44 | (0.12, 1.08) | (0.07, 2.48) |
| 8 | 105 | 0.17 | (0.07, 0.30) | (0.01, 2.94) | 97 | 0.17 | (0.06, 0.30) | (0.01, 2.94) | 8 | 0.45 | (0.13, 0.77) | (0.04, 1.21) |
| 9 | 88 | 0.19 | (0.07, 0.31) | (0.01, 1.89) | 81 | 0.18 | (0.07, 0.30) | (0.01, 0.80) | 7 | 0.47 | (0.20, 1.35) | (0.08, 1.89) |
| 10 | 67 | 0.13 | (0.08, 0.28) | (0.00, 3.40) | 60 | 0.12 | (0.07, 0.25) | (0.00, 0.83) | 7 | 0.39 | (0.15, 1.63) | (0.12, 3.40) |
| 11 | 54 | 0.14 | (0.06, 0.26) | (0.01, 1.35) | 48 | 0.12 | (0.06, 0.25) | (0.01, 1.04) | 6 | 0.65 | (0.35, 0.88) | (0.11, 1.35) |
| 12 | 39 | 0.18 | (0.08, 0.34) | (0.00, 1.59) | 34 | 0.16 | (0.07, 0.29) | (0.00, 0.52) | 5 | 0.86 | (0.36, 1.05) | (0.11, 1.59) |
| 13 | 31 | 0.15 | (0.08, 0.32) | (0.00, 1.37) | 27 | 0.14 | (0.07, 0.25) | (0.00, 0.61) | 4 | 0.43 | (0.22, 0.94) | (0.09, 1.37) |
| 14 | 23 | 0.16 | (0.07, 0.34) | (0.03, 1.10) | 19 | 0.14 | (0.06, 0.22) | (0.03, 0.60) | 4 | 0.75 | (0.39, 1.02) | (0.22, 1.10) |
| 15 | 20 | 0.16 | (0.06, 0.24) | (0.02, 0.92) | 16 | 0.10 | (0.06, 0.19) | (0.02, 0.57) | 4 | 0.75 | (0.47, 0.88) | (0.27, 0.92) |
| 16 | 14 | 0.13 | (0.06, 0.36) | (0.03, 1.10) | 11 | 0.12 | (0.06, 0.16) | (0.03, 0.55) | 3 | 0.69 | (0.44, 1.00) | (0.36, 1.10) |
| 17 | 9 | 0.17 | (0.12, 0.66) | (0.09, 3.93) | 6 | 0.13 | (0.11, 0.17) | (0.09, 0.49) | 3 | 0.98 | (0.66, 3.19) | (0.55, 3.93) |
| 18 | 6 | 0.23 | (0.15, 0.37) | (0.15, 1.50) | 4 | 0.16 | (0.15, 0.27) | (0.15, 0.37) | 2 | 0.90 | - | (0.29, 1.50) |
| 19 | 3 | 1.41 | (0.60, 1.52) | (0.33, 1.55) | 1 | 0.33 | - | - | 2 | 1.48 | - | (1.41, 1.55) |
| 20 | 2 | 0.86 | - | (0.38, 1.34) | 1 | 0.38 | - | - | 1 | 1.34 | - | - |
| 21 | 1 | 0.39 | - | - | 1 | 0.39 | - | - | 0 | - | - | - |
| Pooled | 878 | 0.18 | (0.08, 0.36) | (0.00, 6.19) | 794 | 0.17 | (0.07, 0.32) | (0.00, 6.19) | 84 | 0.67 | (0.22, 1.20) | (0.04, 5.69) |
| **Single dose model** |  |  |  |  |  |  |  |  |  |  |  |  |
| 5 | 154 | 0.21 | (0.09, 0.42) | (0.01, 6.19) | 144 | 0.21 | (0.09, 0.38) | (0.01, 6.19) | 10 | 0.50 | (0.18, 1.69) | (0.07, 3.62) |
| 6 | 138 | 0.19 | (0.10, 0.37) | (0.01, 5.63) | 129 | 0.19 | (0.09, 0.35) | (0.01, 5.53) | 9 | 0.58 | (0.13, 1.39) | (0.11, 5.63) |
| 7 | 124 | 0.20 | (0.07, 0.35) | (0.01, 3.14) | 115 | 0.19 | (0.07, 0.34) | (0.01, 3.14) | 9 | 0.44 | (0.11, 1.08) | (0.05, 2.51) |
| 8 | 105 | 0.17 | (0.07, 0.30) | (0.01, 2.94) | 97 | 0.17 | (0.07, 0.29) | (0.01, 2.94) | 8 | 0.45 | (0.15, 0.78) | (0.02, 1.22) |
| 9 | 88 | 0.18 | (0.07, 0.31) | (0.01, 1.89) | 81 | 0.17 | (0.07, 0.28) | (0.01, 0.80) | 7 | 0.48 | (0.20, 1.35) | (0.09, 1.89) |
| 10 | 67 | 0.14 | (0.07, 0.27) | (0.01, 3.39) | 60 | 0.11 | (0.07, 0.24) | (0.01, 0.83) | 7 | 0.40 | (0.16, 1.64) | (0.11, 3.39) |
| 11 | 54 | 0.14 | (0.06, 0.25) | (0.01, 1.35) | 48 | 0.12 | (0.06, 0.24) | (0.01, 1.04) | 6 | 0.65 | (0.35, 1.02) | (0.10, 1.35) |
| 12 | 39 | 0.15 | (0.08, 0.33) | (0.00, 1.58) | 34 | 0.15 | (0.07, 0.29) | (0.00, 0.48) | 5 | 0.68 | (0.36, 1.06) | (0.11, 1.58) |
| 13 | 31 | 0.15 | (0.07, 0.31) | (0.00, 1.37) | 27 | 0.13 | (0.07, 0.25) | (0.00, 0.60) | 4 | 0.35 | (0.22, 0.87) | (0.09, 1.37) |
| 14 | 23 | 0.16 | (0.08, 0.33) | (0.02, 1.10) | 19 | 0.11 | (0.07, 0.22) | (0.02, 0.58) | 4 | 0.68 | (0.39, 0.96) | (0.22, 1.10) |
| 15 | 20 | 0.15 | (0.06, 0.23) | (0.02, 0.91) | 16 | 0.10 | (0.05, 0.19) | (0.02, 0.55) | 4 | 0.73 | (0.45, 0.88) | (0.27, 0.91) |
| 16 | 14 | 0.13 | (0.05, 0.36) | (0.03, 1.15) | 11 | 0.11 | (0.05, 0.16) | (0.03, 0.53) | 3 | 0.69 | (0.44, 1.03) | (0.36, 1.15) |
| 17 | 9 | 0.17 | (0.12, 0.66) | (0.07, 3.87) | 6 | 0.13 | (0.10, 0.17) | (0.07, 0.46) | 3 | 0.98 | (0.66, 3.15) | (0.55, 3.87) |
| 18 | 6 | 0.23 | (0.15, 0.35) | (0.14, 1.50) | 4 | 0.16 | (0.15, 0.26) | (0.14, 0.35) | 2 | 0.90 | - | (0.29, 1.50) |
| 19 | 3 | 1.39 | (0.58, 1.51) | (0.31, 1.55) | 1 | 0.31 | - | - | 2 | 1.47 | - | (1.39, 1.55) |
| 20 | 2 | 0.84 | - | (0.36, 1.32) | 1 | 0.36 | - | - | 1 | 1.32 | - | - |
| 21 | 1 | 0.37 | - | - | 1 | 0.37 | - | - | 0 | - | - | - |
| Pooled | 878 | 0.18 | (0.08, 0.34) | (0.00, 6.19) | 794 | 0.16 | (0.07, 0.31) | (0.00, 6.19) | 84 | 0.63 | (0.22, 1.19) | (0.02, 5.63) |

